# Forecasting Trajectories of Physiological Mechanics with Sparse Clinical Data Using a Data Assimilation and Machine Learning Hybrid

**DOI:** 10.64898/2026.07.22.26358695

**Authors:** Yanran Wang, J.N. Stroh, Debashis Ghosh, Melike Sirlanci, George Hripcsak, Tellen D. Bennett, D.J. Albers

## Abstract

Clinical decisions for determining optimal patient-specific interventions are complicated prediction tasks that rely on health care professionals’ understanding of physiological mechanisms and their dynamics. These decisions are challenged by (a) observational data sparsity and (b) patient heterogeneity. Here, we focus on estimating and forecasting specific physiological properties—that are not explicitly present in clinical observations—to provide additional features using only data available bedside at the time of decision-making. Mechanistic models of physiological system(s), e.g., physiological ordinary differential equation (ODE) models, provide pathways to compensate for data sparsity by synchronizing the model with observations of an individual patient using data assimilation (DA). However, DA used in a standard computational workflow to estimate constant model parameters from presently-known data is less effective at optimizing state forecasts of the model governed by physiological processes that evolve before new observations are available. Stated simply, we cannot forecast the future evolution of the model because we cannot forecast model parameters. To support next-generation clinical decision support, we develop a new DA and machine learning (ML) hybrid pipeline to estimate and forecast individual future physiological processes by forecasting ODE model parameters. This pipeline overcomes model and DA workflow limitations by stacking a DA-estimated posterior empirical distribution of physiological parameters with longitudinal ML forecasting models. We work within the context of glycemic management in an ICU using EHR data to construct and test a use case. We use synthetic data and real-world clinical data to validate the integrated pipeline and quantify uncertainties.

## 1 Introduction

Healthcare professionals (HCPs) make care decisions by reasoning based on their understanding of the individual patient, clinical practice, health outcomes, pharmacology, and physiological mechanics. These prediction problems become increasingly difficult when available observations of these governing systems [1, 2, 3], e.g., of physiological systems, are extremely sparse [4] and when treatment responses are heterogeneous. Within this HCP-dependent context, predictive models that generate physiological information for *individual patients* can provide additional features that could potentially improve care decisions. To realize this potential, it is imperative that such algorithms model and predict patient-specific information accurately using only data that are available bedside at the time of decision-making. One pathway to providing actionable information is by estimating mechanistic models of physiological system(s) using bedside data available in electronic health record (EHR) [5]. These models [6] are often ordinary differential equations (ODEs) of the form

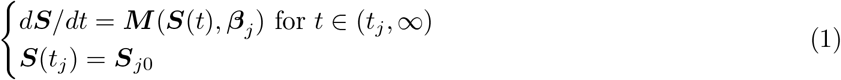

where ***S***(*t*) ∈ ℝ ^*n×*[0,∞)^ are *n* dimensional *model states* or *physiological states* over time domain [0, ∞ ), ***M*** : ℝ ^*n*^↦ ℝ ^*n*^ is a *C*^*r*^ *parameterized function* with *r >* 1 times continuous differentiability, and ***β*** ∈ ℝ ^*m*^ are *m* dimensional constant *model parameters*. Model initialization at an arbitrary starting time *t*_*j*_ requires initial states ***S***_*j*0_ and specified parameters ***β***_*j*_, where *j* ∈ { 1,, *J* } indexes *J* time points. Variables of ***S***(*t*) evolve fast in time, e.g., heart rates, blood pressures, or blood glucose. The parameters, ***β***, are unobservable but represent underlying physiological processes assumed to evolve slowly in time, e.g., vascular plasticity, or insulin reactivity. We assume these parameters evolve more slowly because they are not dynamic in the ODE model and remain fixed when the model is integrated forward in time.

Data assimilation (DA) [7, 8] can be used to synchronize the model states ***S***(*t*) with observations {*y*_*j*_} of an individual patient taken at *J* measurement times 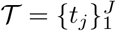 to estimate states and parameters. Further, sequential application can generate joint estimates of initial states ***S***_*j*0_, constant parameters 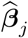 and forecasts of states 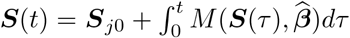 based on these new estimates, for different observed time intervals [*t*_*j*_,*t*_*i*_,+_1_) DA also supports uncertainty quantification (UQ) of estimates and forecasts via ensemble or Markov Chain Monte Carlo methods.

DA computation workflows that include parameter estimation depend on some of the following formulations: filtering that assumes physiological stationarity between time points for point-wise estimation, or smoothing when estimation is computed with several data points over an optimization time window. Either case results in a constant estimator 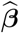 in-between time intervals of estimation instances, as illustrated in Figure 1 (left). However, in many clinical settings, e.g., in an intensive care unit (ICU), patients can be both sparsely measured and highly non-stationary: their physiological systems evolve between measurements, corresponding to a dynamic ***β***(*t*). But, because model parameters are constants in the ODE representations, it is not possible to forecast ***β***(*t*). Instead, model parameters can only change in the model when estimated with new data, e.g., a new measurement *y*_*J*+1_, are available. This gap limits the diagnostic usage of parameter estimates, 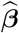, because these constants inferred from presently-known data cannot evolve into the future. Stated simply, although we can forecast model states ***S***(*t*) using parameter estimates up to the current, **we cannot forecast the future evolution of the *model*** because we cannot forecast ***β***(*t*).

**Figure 1:**
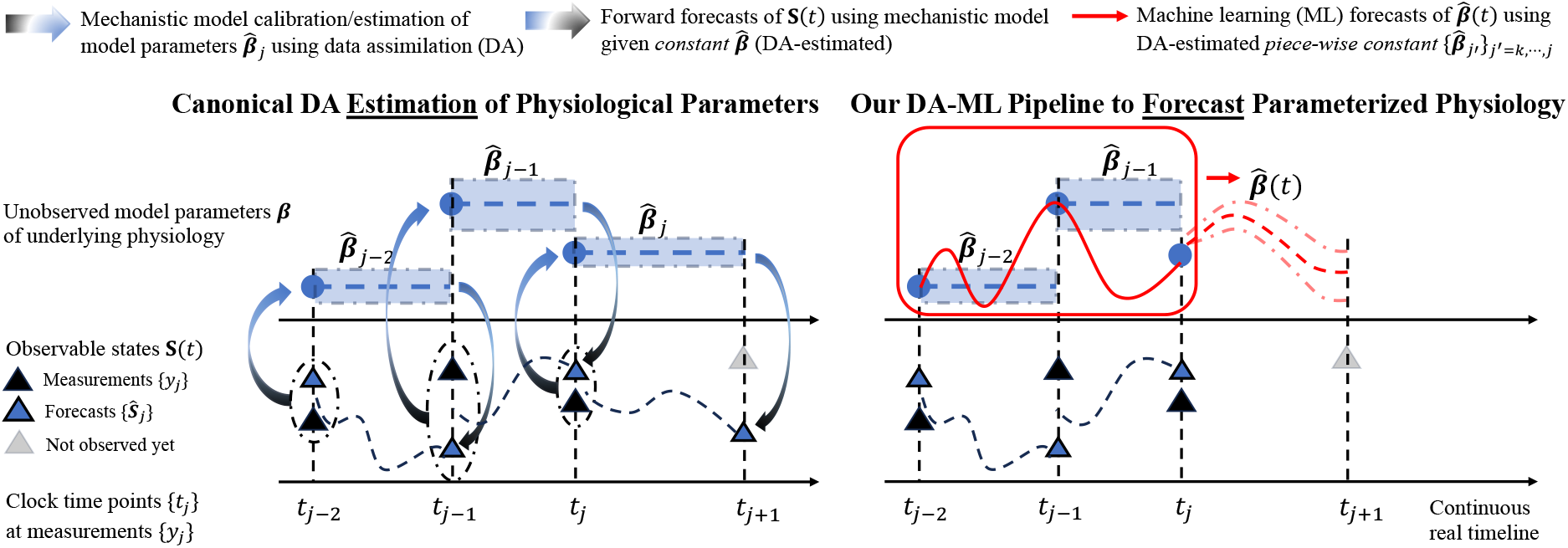
Our ML-DA integrated forecast pipeline (right) vs canonical DA estimation workflow (left). Forecasts 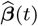 (red dashed curve) are generated from a ML method that uses historical information (in red box) as covariates. These covariates include DA-estimated piece-wise static 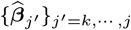 whose linear/nonlinear trajectory curve (red solid curve) is learned under specific parametric assumptions of the ML method.

*Our methodological goal is to develop a new DA and machine learning (ML) hybrid pipeline to estimate and forecast individual future physiological processes—specifically by forecasting ODE model parameters* ***β***_*future*_ *and therefore forecasting the* ODE model*—while addressing the following issues*: (a) observational data sparsity, and (b) patient heterogeneity. To achieve this goal we compose DA and ML in a generalized methods for each individual patient, as illustrated in Figure 1 (right). These ML methods forecast the framework of longitudinal stacking [9, 10, 11] that fills gaps in modeling and DA workflows. We develop a DA-ML integrated pipeline that pushes DA-estimated parameter vectors 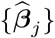 [12] into longitudinal ML model parameters, 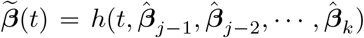, for 1 ≤ *k < j, t*_*j*_ ≤ *t* where the function *h* implicitly incorporates a smoothing step of DA outputs that is necessary because of measurement sparsity and irregularity. Notably, DA estimates properties of physiological processes—here model parameters—that are not directly measured, but this is not the primary development and is based on prior work [13, 12]. Instead, the primary advancement of this paper is the integration of an ML layer to forecast DA-informed ODE models, extending from model calibration with DA to ODE model forecasts with a DA-ML hybrid. This advancement enables forecasting future physiological processes, which cannot be achieved with ML machinery alone because their measurements history is unknowns. In addition, the pipeline propagates uncertainties, enhancing forecast utility by providing HCPs with the estimated reliability needed in clinical decision-making. Our experiments use synthetic data and real-world clinical data to validate the integrated pipeline and to quantify uncertainties in ways standard to UQ [14, 15] and probabilistic ML [16, 17] communities.

To construct and test a use case of our new DA-ML integrated pipeline, we work within the context of glycemic management in an ICU using EHR data available at the bedside. Due to stress dysglycemia or interventions such as drug administration, HCPs regulate patient blood glucose levels with controlled nutrition and insulin therapies according to clinical protocols [18, 19]. The ultradian model of glucose-insulin dynamics [20] used in this paper has only blood glucose as a measured observable with a counterpart state variable *G*(*t*) in states ***S***(*t*). Observational data are very sparse and irregularly sampled: blood glucose measurements are collected nominally once every one to four hours. The model’s primary therapeutic drivers, nutrition (enteral) and exogenous insulin (intravenous, subcutaneous), are also known. Figure 2 (left) shows the model ODEs. Extracting additional physiological information of underlying processes, e.g., insulin reactivity, from these EHR data and providing their forecasts could potentially support decision-making. While establishing that this new source of information will impact clinical decision-making requires careful qualitative analysis and is not investigated here, the potential use of the forecasts we provide is straightforward. Accurate forecasts of model parameters, e.g., insulin reactivity, that control the ODE model choice, and of model states, e.g., blood glucose, have potential uses that range from improved bedside blood glucose management to quantifying patient acuity [21]. In this way, our contribution here represents an effort to enhance the canonical approach by developing a new DA-ML integrated method to forecast individual patent’s underlying physiological system that could provide new, additional, physiologically specific information at the bedside to better inform clinical decision-making.

**Figure 2:**
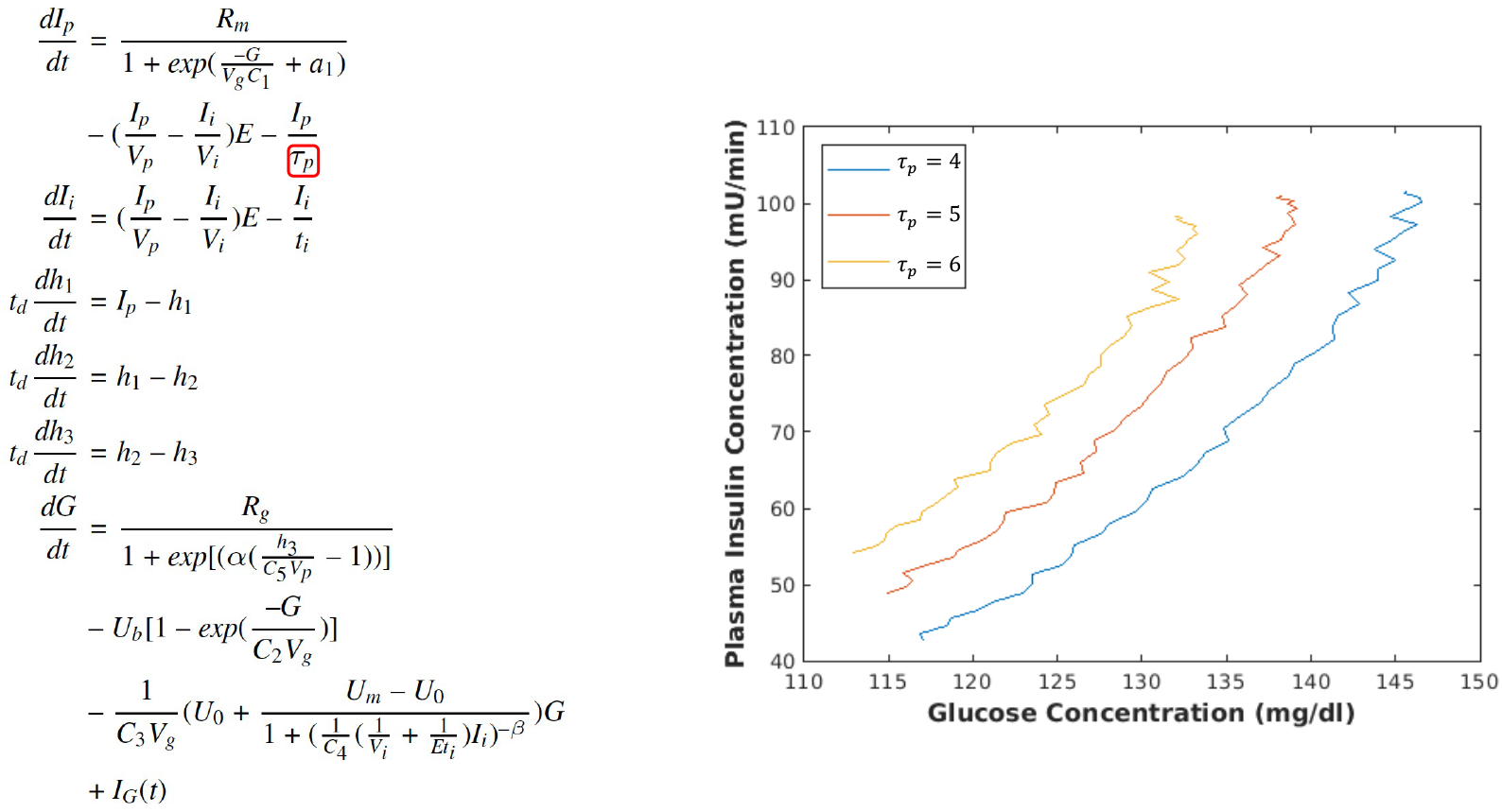
Continuously simulated dynamics (right figure) of blood glucose vs insulin concentration by forward integrating their ODE states (*G, I*_*p*_) and other states (*I*_*i*_, *h*_1_, *h*_2_, *h*_3_) using parameterized functions and specified model parameters of a mechanistic model (left figure, cf. Appendix A.1) that is entirely decoupled from observational data. Three different dynamics are generated from specific *constant* values of scalar physiological parameter *τ*_*p*_. Random measurement noise is added to blood glucose.

## 2 Methods

Our DA-ML hybrid pipeline computes and forecasts trajectories of physiological processes from measurements sparsely collected in time. Data assimilation (DA) is a collection of computational methods for mechanistic model inference and estimation, which links measurement data in the past to contemporary estimates of model states and parameters. This pipeline composes DA with longitudinal models of machine learning (ML) methods to forecast DA-estimated physiological model parameters at future times when no data are yet known. The forecasts of endocrine physiological processes in tube-fed ICU patients serve as a use case. This pipeline has the following four steps: (i) use DA to estimate physiological model parameters from EHR data (Section 2.1), (ii) define moving windows in sequential DA estimation to construct physiological parameter time series (Section 2.2), (iii) smooth parameter trajectories for accurate and robust forecasting (Section 2.3), and (iv) apply ML to forecast these data-informed physiological parameters (Section 2.4). The resulting forecasts of physiological model parameters identify the trajectories of future physiological processes. Additionally, the workflow incorporates uncertainty quantification (UQ) to assess the reliability of forecasts for biological fidelity required by clinical decision-making (Section 2.5).

### 2.1 Step One: Characterizing Individual Time-specific Physiological Processes with DA-driven Physiological Parameter Estimation

The ML-DA hybrid pipeline step one is to characterize time-specific physiological states of individual patients using data assimilation (DA). This is achieved by estimating physiological parameters ***β*** of a mechanistic ODE model from observational data over a fixed time window, data assimilation window (DAW). The result is a vector of computational biomarkers of unmeasured or unmeasurable physiological processes. Our previous work [12] includes a full explanation of this methodology.

#### 2.1.1 Ultradian Physiological ODE Model Represents Glucose-Insulin Dynamics

The ultradian model [20]—a set of six ODEs with 21 parameters—approximates oscillatory dynamics of glucose-insulin mechanisms [22] at time scales relevant for ICU glycemic management. This relatively simple and widely validated model, entirely decoupled from observational data, embeds knowledge of essential physiological features. This model *d****S****/dt* = ***M*** (***S***(*t*), ***β***) comprises three components: *physiological states* ***S***(*t*) = (*G*(*t*), *I*_*p*_(*t*), *I*_*i*_(*t*), *h*_1_(*t*), *h*_2_(*t*), *h*_3_(*t*)) ∈ ℝ ^6^, continuously differentiable *parameterized functions* ***M***, and constant *model parameters* ***β***. The mathematical formulation and details of this model are included in Appendix A.1. Blood glucose, which has its observable model counterpart *G*(*t*), is measured in the ICU and managed by controlling nutrition and exogenous insulin (source of plasma insulin *I*_*p*_ and interstitial insulin *I*_*i*_), while other quantities modeled in ***S***(*t*) are typically not observed. These parameterized functions *M* frame mathematical hypotheses about physiological and biochemical processes constraints that relate physical sub-processes to ***S***(*t*). Synchronizing this continuous-time state-space map with the present individual data injects these personalized physiological hypotheses, enabling the extraction of mechanistic knowledge from observations. Model parameters ***β*** define underlying physiological processes and specify a model representation of an individual physiological system. For example, time constant of plasma insulin clearance timescale (*τ*_*p*_) in equation *dI*_*p*_*/dt* nonlinearly affects the relationship between plasma insulin concentration and glucose concentration (Figure 2 (right)). Identifying the optimal model via inferred model parameters leads to characterizing the physiological states of individual patients.

#### 2.1.2 DA Estimates Model Parameters to Characterize Physiological Sub-processes

Data assimilation (DA) [23] is a set of computational techniques [24, 25, 26, 24, 27] developed and applied to mechanistic models, e.g., in weather forecasts [25][24], moon landing [7], aerospace [8], and robotics [28], and medicine [29][23]. To estimate model parameters ***β*** that specify individual physiological processes [30, 31], this work uses DA within the Bayesian inverse problem context [32, 33, 34] in its smoother formulation [12] that borrows individual information across multiple time points in a time window, data assimilation window (DAW). This model calibration procedure [35] involves stochastic optimization of model parameters ***β*** that uses a Metropolis-Hastings-within-Gibbs of Markov chain Monte Carlo (MCMC) algorithm [36]. Operationally each model parameter in ***β*** is estimated using 10 Markov chains with different initial conditions, where each Markov chain comprises 20K sampling iterations. Extra care is taken to qualify numerically generated solutions of inferred parameters. The final 20% of sampled iterations are retained in qualified Markov chains that are screened by convergence and accuracy criteria [12], including mean squared error (MSE) minimization and the Geweke statistic. The combined iterations 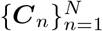 with *N* sampled data points empirically approximate the parameters posterior probability distribution ℙ (***β***|*y*) that minimizes the mean squared error (MSE) between model-estimated states and measurements {*y*_*j*_ }_*j*∈*J*_ at times { *t*_*j*_ } _*j*∈*J*_ for *J*= { 0,, *J* }. In our application, these measurements correspond to blood glucose that has its modeled counterpart *G*(*t*) in states ***S***(*t*), while nutrition and exogenous insulin data that are required forward model inputs are not used in model calibration. This DA estimation results in time-specific, continuous-distributed model parameters 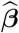 that characterize individual’s underlying physiological states.

#### 2.1.3 Parameter Selection for Model Calibration and Identifiability

Estimating all parameters of the ultradian model [20] is likely unidentifiable [23, 37, 38] given observational data sparsity and parameter covariance. Even with sufficient data, adjusting parameters to fit the model essentially involves a nonlinear optimization problem that may have multiple solutions [39]. This model parameter calibration requires selecting parameters that balance model identifiability, flexibility, and clinical usefulness [12], while excluded parameters are set at their nominal values. The selected parameters ***β*** = (*a*_1_, *τ*_*p*_, *R*_*g*_) ∈ ℝ ^3^ of insulin reactivity correspond to pancreatic insulin secretion (*a*_1_), insulin degradation via liver and kidney (*τ*_*p*_), and insulin resistance (*R*_*g*_). System identification through this parameter subset and estimation approach (cf. Sec. 2.1.2) are detailed in previous work [37, 12]. Inference of this parameter triplet characterizes three underlying physiological processes that are important in the hepatic-renal-insulin subsystem.

### 2.2 Step Two: Constructing Time Series of Physiological Parameters to Characterize the Evolving Physiological System

The pipeline step two quantifies slow changes in physiological processes by constructing time series of static model parameters estimated on *W* moving windows of individual patient data. These overlapping data assimilation windows, or DAWs {*D*_*w*_ } _*w*∈*W*_ indexed and ordered by *W* = {1,, *W* }, are chosen such that the physiological system is roughly stationary and model identifiability is managed within each *D*_*w*_ estimates parameters ***β*** over time interval *D*_*w*_ based on *Y*_*w*_ for an indexed time series {***β***_*w*_ }_*w*∈*W*_ that while accounting for irregularity of observation times. We define these DAWs *D*_*w*_ = [(*w* − 1)*F*, (*w* − 1)*F* + *L*] to have a width *L* = *aδt* (in hours) and forward shift *F* = *bδt* (in hours), where width and shift parameters *a, b* ∈ Z^+^ with *a > b* and fixed time step *δt >* 0. This construction results in sequential time intervals of equally-weighted observational data sets *Y*_*w*_ = { *y*_*j*_ | *t*_*j*_ ∈ *D*_*w*_ } similar to the application of rectangular window functions used in signal processing [40, 41]. Sequentially DA estimates parameters β over time interval Dw based on Yw for an indexed time series 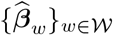 that quantifies the trajectory of physiological processes. Figure 3 shows an example of constructing and estimating on two sequential DAWs.

**Figure 3:**
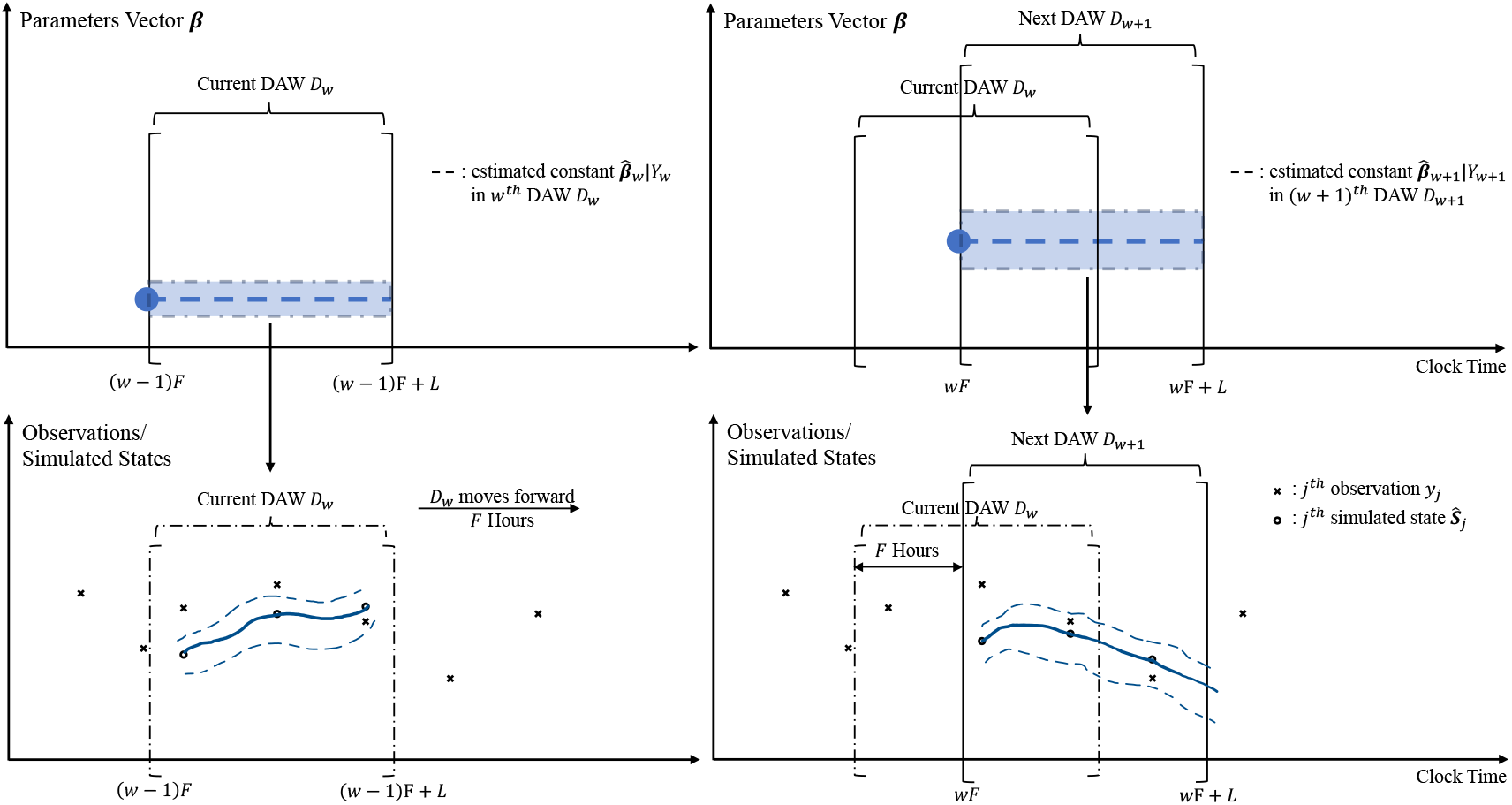
Illustration of constructing time series of physiological parameters 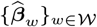 using two sequential data assimilation windows (DAWs) of fixed lengths *L*, where the next DAW, *D*_*w*+1_, shifts forward from the current DAW, *D*_*w*_, by *F* hours. Physiological model parameters in each DAW *D*_*w*_ are estimated by synchronizing the model with glucose measurements *Y*_*w*_ = {*y*_*j*_ |*t*_*j*_ ∈ *D*_*w*_} (bottom row cross signs). Bounded by 95% CIs in dashed lines, solid lines refer to the average of model-simulated state *S*_*j*_ related to blood glucose at each measurement time points *t*_*j*_ (bottom row circles). The assumed stationary physiological system is represented by constant parameters 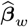 and 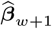 (top row) within *D*_*w*_ and *D*_*w*+1_.

### 2.3 Step Three: Smoothing Overlapping DA Estimates to Improve ML-Driven Forecasting Reliability

The pipeline step three smooths the time series of DA-estimated stationary parameters 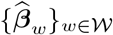 using a moving average weighted by overlap proportion to increase ML forecasting robustness and reliability. According to Eq. (2),

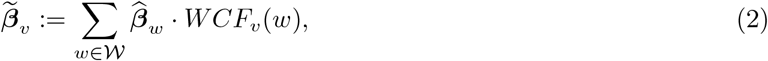

we generate a sequence of smoothed analysis windows (SAWs) defined by *S*_*v*_ := [(*v* − 1)*F, vF* ], *v* ∈ *W* to describe the DAWs {*D*_*w*_}_*w*∈*W*_ overlap on which we continue to assume the physiological system is stationary. Importantly, each estimate 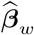 also represents the uncertainty of parameter distribution approximated by MCMC samples for an individual patient. As such, the uncertainties of approximated parameter trajectories from step two are also propagated in this step to support the whole pipeline UQ. This smoothing process weights the DAW estimates in proportion to their relative representation in *S*_*v*_ using function *WCF*_*v*_(*w*) formulated by

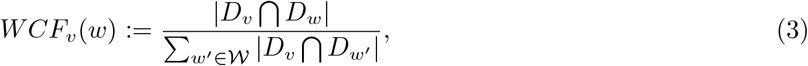

which is illustrated in Figure 4 ). Here notation |*D*_*v*_ ∩ *D*_*w*_| represents the length of overlap between intervals *D*_*v*_ and *D*_*w*_. This step lessens the impact of data variability and sparsity (*e*.*g*., most *L*-hour DAWs have fewer than *L* data points) and reduces overlapping DAW estimates to a smoothed time series with finer stationary timescale *F* . Note that SAWs and DAWs have the same index set *W*. The results, 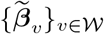, are longitudinal vectors of smoothed parameters that represent slowly evolving physiological processes and will become covariates of the ML machinery to forecast these processes.

**Figure 4:**
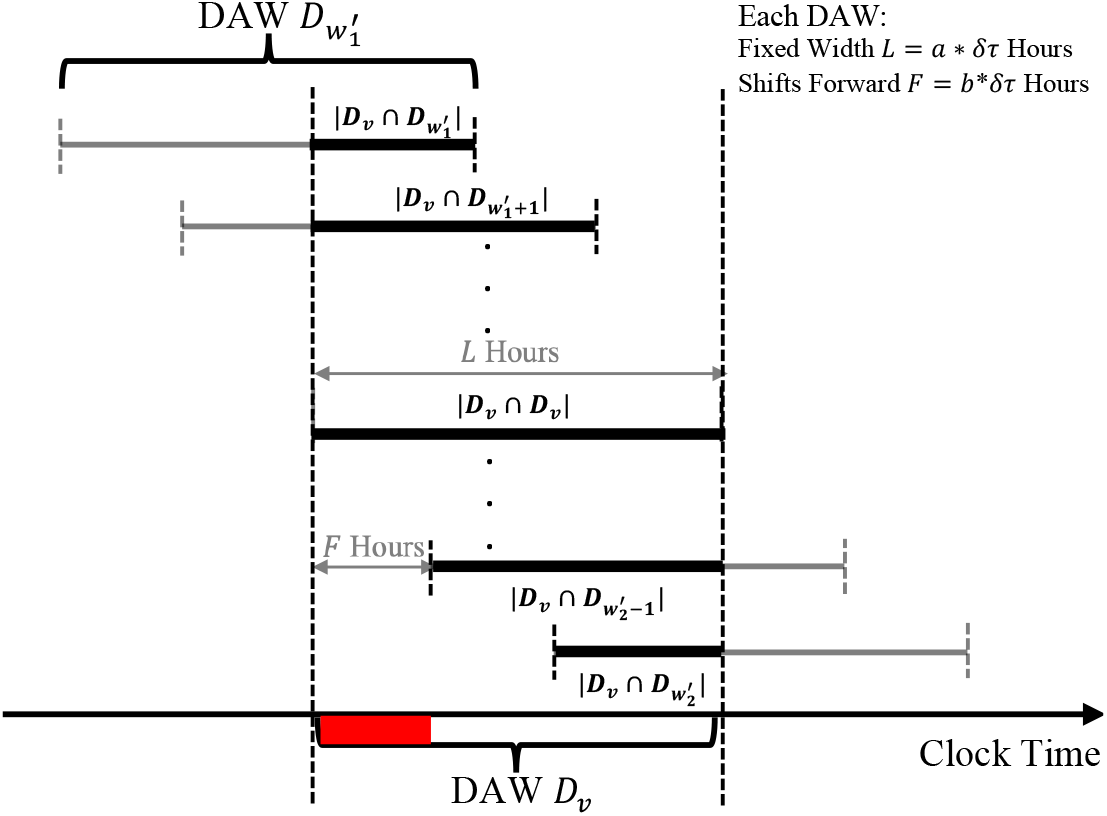
Illustration of weight coefficient function *WCF*_*v*_(*w*) based on surrounding {*D*_*w*_}_*w*∈*W*_ overlapped with *D*_*v*_ for calculating one specific SAW-indexed estimate, 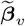, highlighted in red. The range of DAWs with nonzero weights are determined by ratio 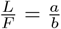 and *v*. Each SAW interval starts at one DAW and ends the start of the next.

#### 2.3.1. Evaluate Hyperparameter of Inference Timescale

We choose SAW hyperparameter *a* to address overestimated variation from frequent windowed estimation using sparse observational data, while setting explicit hyperparameters *b* and short time interval *δt* as constants. This parameter *a* determines stationary inference timescale *L* and controls properties of 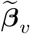 such as 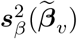, which is the point-wise temporal variability of the smoothed sequential parameter estimates. An optimal *a* balances variance over time, *V ar*_*v*∈*W*_ (E_***β***_(***β***_*v*_ )), and DAW data sufficiency (to maximize *a*) with temporal resolution and noise reduction (to minimize *a*). This evaluation process, quantitatively and qualitatively, is similar to finding a minimal sufficient statistic that balances sufficient statistical representation with temporal resolution to maintain all information required for statistical inference. For methodological simplicity, a common value of *a* that we adopt for all patients is manually selected by analyzing for several individuals as illustrated in Figure 5.

**Figure 5:**
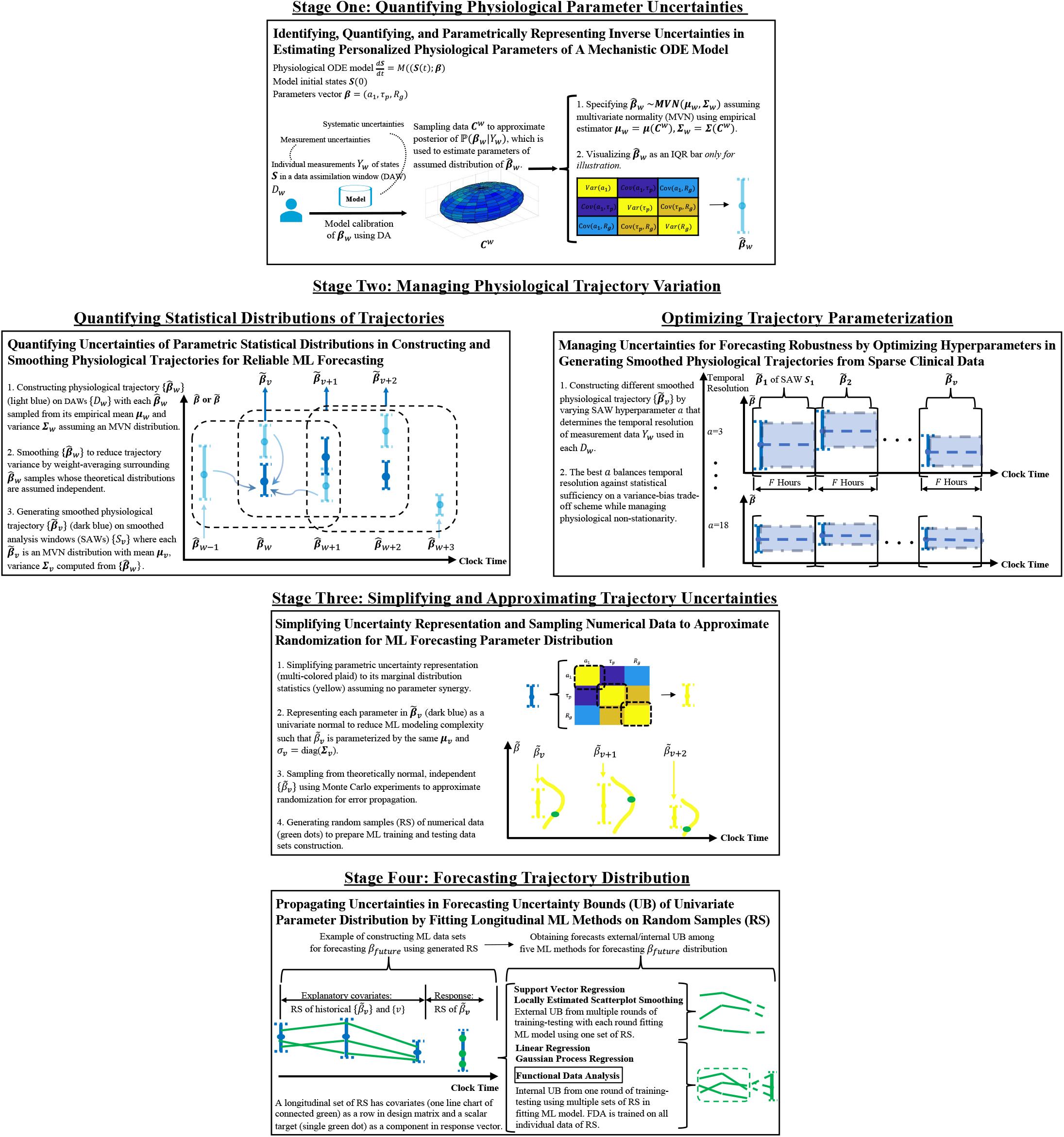
Overall pipeline workflow of uncertainty identification, quantification, management, and propagation, separated into four stages.

### 2.5 Step Four: Forecasting Individual Slowly-Evolving Physiological Processes Using ML Models of Longitudinal Regression

The pipeline step four applies longitudinal ML methods to smoothed parameters sequences, 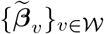, to forecast physiological processes ***β***_*future*_. The temporal forecast resolution depends on the DAW-SAW windowing-smoothing timescales in steps two and three. The longitudinal data constrains the complexity of models that we can use in the analysis. As such, we formulate univariate parameter forecasts using five ML algorithms (ordered by increasing flexibility): linear regression, support vector regression, Gaussian Process regression, locally estimated scatterplot smoothing, and functional data analysis. Depending on each ML method’s limitation in managing uncertainties and trajectory variation reconstruction, these methods are applied over the population estimates to generate continuous ***β***_*future*_ as discussed below, although only functional data analysis is trained on population level. We do not consider a persistent model (***β***_*v*+1_ ∼ ***β***_*v*_ ) that assumes stationarity between measurement points. While this model will allow reasonable forecasts for a short time in the case of dense data and slow changes, it is known to be inaccurate for any forecast time horizon in the case of sparse data and rapid changes in health states and interventions.

#### 2.4.1 Construction of Training and Testing Data Sets for Longitudinal ML Models

To emphasize the application to brutally sparse and irregular data, we focus on the univariate forecasting problem by ignoring parameter synergy. This simplification potentially limits the forecast complexity and reduces error by not regularizing over both space and time while simultaneously estimating covariance evolution. The longitudinal physiological data for a specific patient *i* in the univariate parameter case are the sequential vectors

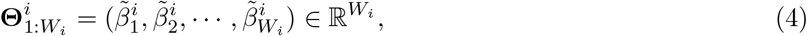

where 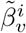 is single parameter component of SAW-indexed estimate 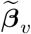 of patient *i*, and *W*_*i*_ indexes the windowing set *W*_*i*_. We assemble the training data set for patient *i* as

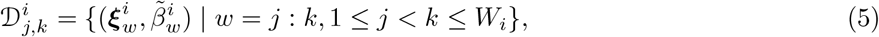

where *j* : *k* is SAW index span of each training data element, 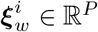 is *P* -dimensional individual’s feature vectors, 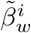 is the associated target. These longitudinal vectors of limited parameter history prior to SAW *S*_*w*_, 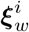, consists of historical SAW indices {*w′*} and/or SAW-indexed estimates 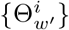 for *w′* = *w*−*P* : *w*−1 ⊆ *W*_*i*_. The default restriction of 1 *< P < j* ensures we only use previously generated information as training features. The accordingly constructed testing data set to evaluate forecasts, for a SAW *S*_*v*_′with index *k < v′* ≤ *W*_*i*_, has 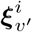, as its input feature vector and 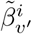, as the associated target. The ML forecast problem of estimating an individual’s *w*th history in time interval *S*_*w*_ from these data is

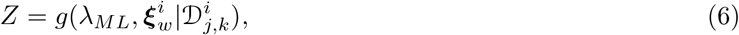

where *g* refers to the general ML function or algorithm and *λ*_*ML*_ is the collection of ML hyperparameters. For notation simplicity, we aggregate the training feature vectors 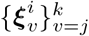 into rows of design matrix 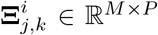 with *M* = *k* − *j* + 1, and similarly, 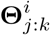 is the vector of training scalar responses 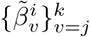. Further mathematical details of design matrix construction are included in Appendix A.2. We present the specific forms of *g* and *λ*_*ML*_ for each machinery in Appendix A.3. These forms exclude linear regression because of its close similarity to locally estimated scatterplot smoothing in the linear case.

### 2.5 Uncertainty Identification, Quantification, Management, and Propagation

Probabilistic representation of uncertainties [14, 15] towards trustworthy uncertainty forecasts [23] is essential for clinical utility [14, 42, 23] but no universally accepted framework exists in many settings [43]. With this in mind, we identify, quantify, manage, and propagate uncertainty through ML-DA hybrid pipeline in four stages discussed below, as illustrated in Figure 5 . To build a clearer connection between UQ and its presence in pipeline step-wise development, we point out the correspondence between these UQ stages (one to four) to the pipeline steps (one to four).

#### Stage One of Figure 5

UQ stage one estimates the inverse uncertainties [42] of three physiological parameters ***β*** = (*a*_1_, *τ*_*p*_, *R*_*g*_). These parameters are specified in a mechanistic ODE model, *d****S****/dt* = ***M*** (***S***(*t*), ***β***), of model states ***S***(*t*) and parameterized function *M* using individual observational data *Y*_*w*_ of a patient. Based on predictive estimation theory in UQ [15], UQ stage one incorporates uncertainties from (i) probabilistic measurement error in observational data, and (ii) non-probabilistic potential systematic uncertainties due to specification errors or model discrepancies. These uncertainties are reflected in estimating time-specific physiological parameters ***β***_*w*_ within a data assimilation window (DAW) *D*_*w*_ indexed by *w*, where estimation bias is reduced from keeping only data-optimized samples. Specifically, we approximate the parameter posterior probability measure P(***β***_*w*_ |*Y*_*w*_) using an empirical distribution function (EDF) formed by combined MCMC samples 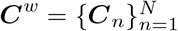 with *N* data points. For ease of pipeline implementation limited by computing resources, we further parametrically represent the approximated distribution, 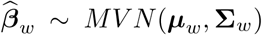, as a multivariate normal (MVN) distribution with a mean vector ***µ***_*w*_ = E_*N*_ [***C***^*w*^] and covariance matrix **Σ**_*w*_ = E_*N*_ [(***C***^*w*^− ***µ***_*w*_ )(***C***^*w*^ − ***µ***_*w*_ )^*T*^ ] according to the empirical rule under MVN assumption. UQ stage one corresponds to pipeline step one (cf. Sec. 2.1).

#### Stage Two of Figure 5

UQ stage two constructs the trajectories of DAW-indexed physiological parameters, 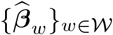, by estimating ***β*** on a sequence of forward-shifted, fixed-width time intervals DAWs, {*D*_*w*_}_*w*∈*W*_, with indices set *W* = {1, · · ·, *W* }. According to UQ stage one, the DAW-specified random variable, 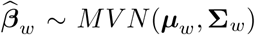, follows a MVN distribution with empirical mean ***µ***_*w*_ and covariance **Σ**_*w*_. Note that physiological parameter estimates implicitly incorporate observation densities of their associated DAWs; for example, sparse data lead to wider posterior distributions and thus higher quantified parameter uncertainties. For explicit UQ, we also assume temporal independence between any estimation *D*_*w*_ because ML regression methods generally prefer independent covariates to estimate uncertainties. Further, the use of smoothed analysis windows (SAWs), *S*_*v v*∈*W*_, allows balancing physiological non-stationarity with data sparsity in constructed trajectories from overlapping parameter estimates. The smoothed physiological trajectories, 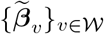, consist of SAW-indexed, MVN-distributed, physiological parameters, 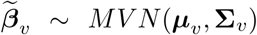, by averaging surrounding independent, MVN-distributed DAWs. Importantly, parameter smoothing weights the DAWs relative to their intersections with a given SAW, implicitly reducing the influence of temporal dependence among estimates from overlapping DAWs on the smoothed estimates. Analogous to a sensitivity analysis, we choose SAW hyperparameter *a* to balance temporal resolution with the statistical sufficiency needed to resolve essential variability over time, *V ar*_*v*∈*W*_ (E_***β***_(***β***_*v*_ )), while simultaneously managing point-wise temporal variability in **Σ**_*v*_ which is 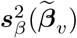. UQ stage two corresponds to an operational combination of pipeline steps two and three (cf. Sec. 2.2, 2.3).

#### Stage Three of Figure 5

UQ stage three systematically simplifies uncertainty representations and approximates randomization for forecasting univariate physiological parameter. Extending from previous distributional assumptions, each MVN representation, 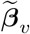, is simplified to its marginal univariate normal by ignoring off-diagonal elements in **Σ**_*v*_. As such, 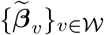 is reduced to a scalar time series, 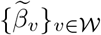, with mean ***µ***_*v*_ and variance 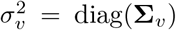. To approximate randomization for error propagation [44], numerical data are generated from the simplified parametric quantities using Monte Carlo experiments. In each experiment of random numbers [45], a longitudinal set of random samples of 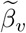 according to SAWs ordering is simultaneously drawn from each of their univariate normal distribution. Essentially, multiple sets of random samples from a collection of these experiments form an EDF that approximates the measure of non-parametric joint probability in UQ stage one. By generating one or multiple sets of random samples, UQ stage three is the preliminary process for constructing the design matrices and response vectors in ML training and testing data sets. UQ stage three corresponds to a transition between pipeline steps three and four (cf. Sec. 2.3, 2.4).

#### Stage Four of Figure 5

Our quantity of interest is the distribution of univariate physiological parameter forecast *β*_*future*_ at future time points. This is because, besides a point forecast, we are also concerned about forecast uncertainty bound (UB)—the uncertainties of uncertainty [43]—that provides the distribution of forecasting values for evaluation. Conceptually, UQ stage four forecasts the distribution of univariate physiological parameter using historical numerical data—an interval, histogram, or quantile—that represent the parametric EDF of the same physiological parameter. Resembling the uncertainty propagation process [15], UQ stage four further justifies how constructing ML data sets from random samples impacts uncertainty propagation through ML algorithms in a way that influences forecast distribution while accounting for errors.

Given the flexibility in generating numerical data in design matrices and response vectors of ML data sets, we can choose either (i) a ‘fixed’ value to simplify experimental design but with less accuracy, e.g., only one random sample or only the mean value, or (ii) ‘multiple’ observations as in observational studies, e.g., multiple sets of random samples. This flexibility renders a permutation of four combinations as rows in Table 1, while the usefulness of each combination depends on how ML algorithms generate forecast UB in the uncertainty propagation process. Here we introduce five ML forecasting methods that are separated into two groups according to their algorithmic constraints. Two algorithms, including support vector regression and locally estimated scatterplot smoothing, have no direct modeling of randomness hence need external forecast UQ based on repeated random sampling. In each training-testing round, we fit these models using one random sample to output a point forecast such that we obtain the forecast UB after several rounds. We also justify involving multiple random samples in one training-testing round when using other three algorithms that have explicit internal mechanism for forecast UQ, including individual-trained linear regression and Gaussian Process regression, and population-trained functional data analysis. The necessity and consequence of using each combination of random samples varied by ML algorithms are delineated in Table 1 . For methodological consistency, though, using multiple random samples for both design matrices and response vectors is the only combination appropriate for all ML algorithms to produce forecasting UB. UQ stage four corresponds to pipeline step four (cf. Sec. 2.4).

**Table 1:** The impact of constructing ML data sets on generating forecast uncertainty bounds (UB), relative to the forecasting power (✓, **X**) and randomness modeling capacity of each ML algorithm. Establishing the necessity of involving multiple random samples (RS) in both design matrices and response vectors of ML data sets, among all four combinations (rows) from the permutation of {‘Fixed’,’RS’} x {‘Fixed’,’RS’}.

| Combination | Forecasting | Linear Regression | Support Vector Regression | Locally Estimated Scatterplot Smoothing | Gaussian Process Regression | Functional Data Analysis (population-trained) |
| --- | --- | --- | --- | --- | --- | --- |
| Fixed x Fixed | parameter mean | ✓; but forecast UB does not represent propagated uncertainties in each SAW | ✓; but no forecast UB | ✓; but no forecast UB | ✓; but forecast UB does not represent propagated uncertainties in each SAW | ✓; but forecast UB only represents propagated uncertainties at population instead of individual level |
| RS x Fixed | parameter mean | ✓; but removes linear dependence hypothesis of response data | ✓; but narrower forecast UB | $\checkmark\checkmark$ preferred because of stability | **X** requires response data variability | ✓ although has higher complexity in spline-smoothing RS because of using concurrent variables fitted in functional principal analysis. |
| Fixed x RS | parameter distribution | ✓; but could overestimate variance | $\checkmark\checkmark$ wider forecast UB | ✓; but less stable | **X** only works in theory and also produces constant forecasts for different SAW due to unity in covariance function | ✓ although less stable with generalized least square estimates of spline coefficients due to collinearity in design matrix. |
| RS x RS | parameter distribution | $\checkmark\checkmark$ ideal | $\checkmark\checkmark$ ideal | ✓ stable although forecast UB is wide | ✓ although requires avoiding constant inputs from time covariates by adding 'variability' | ✓ although has complex overlay effect of RS x Fixed and Fixed x RS. |

## 3 Validation and Performance Evaluation

The ML-DA hybrid pipeline we develop to estimate and forecast trajectories of physiological mechanics with sparse clinical data has four steps, each of which we validate or evaluate. We evaluate each step of the pipeline in two ways. First, we validate and evaluate pipeline ability to forecast physiological parameter estimation for a simulation study where we know the ground truth. Second, we evaluate pipeline using real-world clinical data.

The pipeline step one we develop in this paper is itself a pipeline to estimate constant physiological parameters with DA developed and validated in our prior work [12] in the same physiological context and with the same data, and so we will not validate this step here. This step provides the raw information we will use in the remaining three steps of our pipeline here. These remaining steps are validated and evaluated both qualitatively and quantitatively by comparing the ML forecasts of parameters with their ground-truth in the case of the simulation study, or with their estimated values in the case of the clinical data study.

### 3.1 Simulation Study

The simulation study has four steps. *First*, we define the ground truth for model parameter trajectories of ultradian model [20] to represent the slowly evolving physiological processes. Our prediction task will be to estimate and then forecast these processes represented by model parameters. The model parameters we focus on are ***β*** = (*a*_1_, *τ*_*p*_, *R*_*g*_) where *a*_1_ corresponds to the glucose concentration for which insulin is secreted, *τ*_*p*_ represents a time constant of plasma insulin clearance timescale, and *R*_*g*_ is maximum rate of delayed insulin-dependent glucose production. In the setting we are studying, real clinical data are so sparse that estimating the joint distribution of the parameter vector and its evolution in time is not possible. Instead, we estimate each parameter and its trajectory as a univariate process. Accordingly, we define the ground truth for and estimate model parameter trajectories independently. *Second*, we create synthetic clinical data by integrating the ultradian model forward in time with prescribed parameter variations and external nutrition administration over a seven-day period. This creates a glucose time series that can be subsampled to resemble what is present in ICU data, e.g., continuous glucose monitor (CGM) data, for the purpose of pipeline evaluation. Finally, Gaussian noise is added to these time series. *Third*, we apply the estimation and forecasting pipeline to these simulated glucose data to demonstrate the pipeline’s ability to reconstruct the parameter trajectories and their uncertainties. We generate these estimates from individual simulations of patient data, without borrowing data across patients. Forecasts of these personalized parameters are generated with ML in the form of longitudinal regressions using various methods. And *Fourth*, we evaluate ML forecasts of individual insulin parameter’s distribution by computing the root mean squared error (RMSE) between the ML forecasts and the ground truth of pre-prescribed parameter variations.

#### 3.1.1 Generating Synthetic Clinical Data

We perform two simulated experiments (A, B) that resemble real-world scenarios of glucose-insulin dynamics driven by unobserved external factors. Experiment A assumes physiological changes that occur as step functions changing from/to baselines to mimic rapid and immediate response to an intervention effect such as drug administration. For experiment A the physiological change is represented by a model parameter in the simulated data changing according to a step function. Experiment B is designed to mimic slower physiological changes that onset and then decay continuously to baselines. For experiment B the physiological change is represented by a model parameter in the simulated data changing with a functional shape defined by the chi-squared distribution with df=4. We set these baselines as nominal values of physiological parameters (cf. Appendix A.1). The synthetic clinical data generated from these simulations and other necessary clinical data we set are restricted to the variables observed in the clinical setting, here blood glucose, nutrition administered, and exogenous insulin. To simplify these analyses we set nutrition to be constant and no exogeneous insulin administration. To generate sparse measurement data, we first generate dense data by integrating the model forward at a time resolution of one point per minute. Then we down sample these simulations to five-minute measurements to simulate a CGM. Finally, we add Gaussian noise with mean zero and variance 25 (mg/dl) to each glucose measurement.

#### 3.1.2 Qualitative Physiological Parameter Trajectory Evaluation

Figure 6 demonstrates the pipeline’s estimation and forecasts of the trajectory of insulin clearance (*τ*_*p*_) parameter for experiments (A, B) using three ML methods: linear regression, support vector regression, and Gaussian process regression. This figure shows qualitative accuracy, variability, and uncertainty of the physiological parameter estimates and forecasts of parameters as they are changing in time for varied time horizons. While all the methods produce reasonable forecasts, qualitatively linear regression appears to provide the most accurate forecasts. In supplementary materials we include six animated videos for four ML methods dynamical forecasts of all three insulin parameters under both experiments.

**Figure 6:**
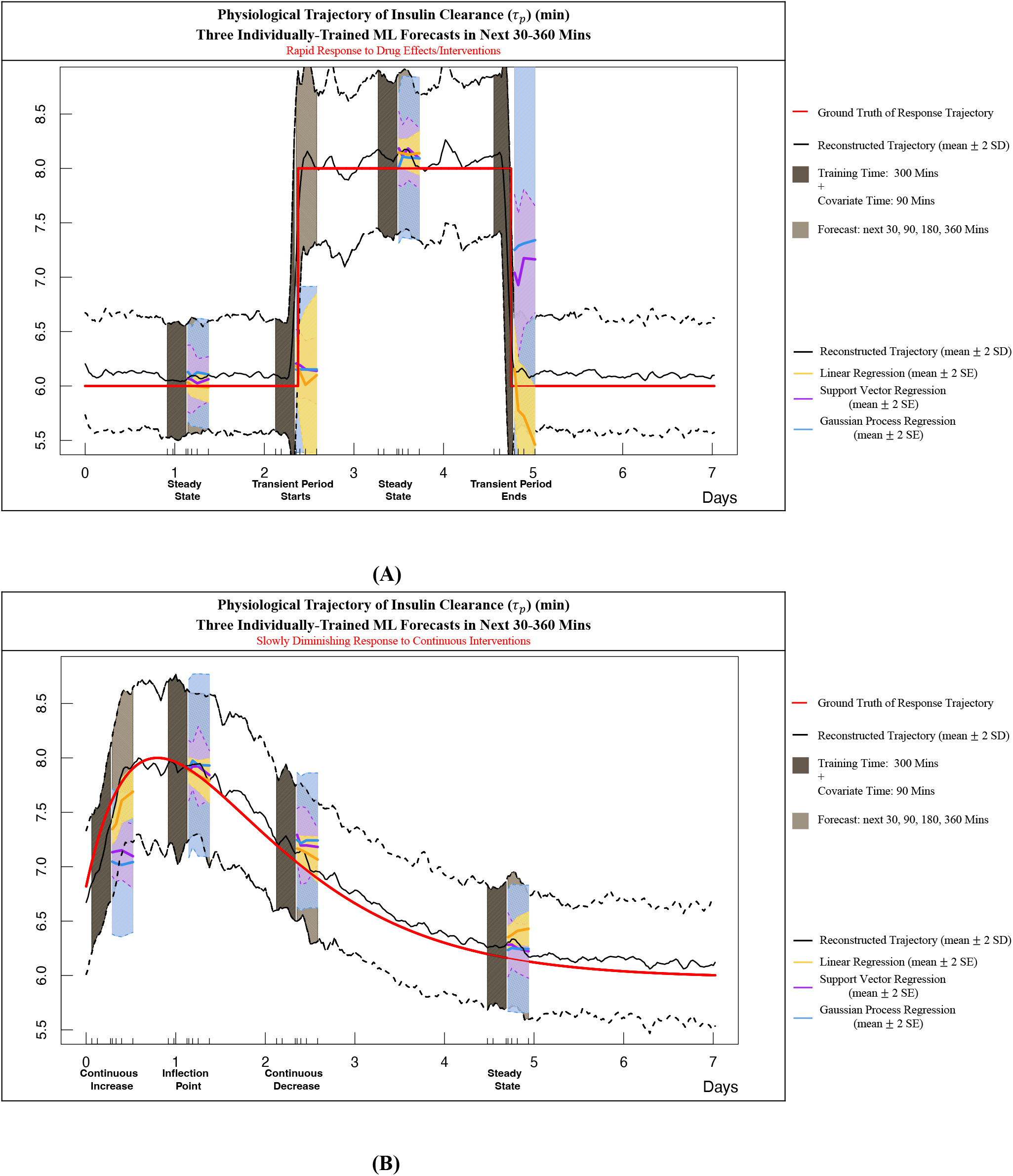
Reconstructing parameter trajectory of insulin clearance (*τ*_*p*_) using one simulation of glucose based on assumed physiological responses that are (A) rapid and immediate, or (B) slower with onset and decay continuously. Primary results contrasting three ML methods forecasted mean (yellow, purple, blue solid curves) are revealed by their accuracy to be close to ground truth (red curve) and their adaptability to capture variability (yellow, purple, blue dashed curves). This reconstruction depends on explicit hyperparameters *b* = 1, *δt* = 30 minutes and chosen inference hyperparameter *a* = 4. Using ML data sets generated from reconstructed trajectory, three individually-trained ML forecasting methods take the past 300 minutes as training data with each data point containing features of the past 90 minutes. Forecasts provided by locally estimated scatterplot smoothing are excluded for better visualization.

#### 3.1.3 Quantitative Physiological Parameter Trajectory Evaluation

To quantitatively evaluate the pipeline to compare ML forecast methods, we construct 10 simulations for each experiment-parameter tuple formed by elements from simulation experiments (A, B) and physiological parameters ***β*** = (*a*_1_, *τ*_*p*_, *R*_*g*_). Meaning, this construction results in 60 total simulated clinical situations (10 simulations * two experiments * three parameters). In each simulation the nutrition intake and size of assumed parameter changes vary randomly by *±* 20% relative to the baseline values of the parameters. In this way, each one of 60 simulations represents the glucose-insulin dynamics of an individual ICU patient with varied insulin responses given constant nutrition. Table 2 shows the forecast accuracy characterized by the root mean squared error (RMSE) between the ML forecasts and the ground truth of model physiological parameter trajectories over 30-360 minutes. Additionally, we report bias and forecasts interval coverage probabilities with their uncertainties in Table A.1 and Table A.2 . Overall linear regression and support vector regression have the lowest average RMSE and bias, while Gaussian process regression and locally estimated scatterplot smoothing (LOESS) have the highest coverage probability. LOESS has the largest average RMSE and bias with extremely high errors, making its forecasts the least reliable despite of high coverage probability.

**Table 2:** Evaluation of ML forecasting univariate physiological parameter distributions using root mean squared error (RMSE) averaged over 10 simulations with uncertainty bounds (*±* SE). This evaluation process is separated by experiments assuming insulin response that are (A) rapid and immediate, or (B) slower with onset and decay continuously, with forecast accuracy of parameters (row sections) at four future time points (columns). For each parameter, cells highlighted in color blue have the lowest average RMSE among four ML methods. According to our UQ approach (cf. Sec. 2.5), forecasting parameter distribution is based on 20 random samples (RS) when using linear regression (LR), support vector regression (SVR), and locally estimated scatterplot smoothing (LOESS). Forecasts are based on 10 RS for faster computation when using Gaussian Process regression (GPR).

| Accuracy of Forecasting Underlying Physiological Processes Using Average RMSE ± SE | | | | | | | | | |
| --- | --- | --- | --- | --- | --- | --- | --- | --- | --- |
| Physiological Parameters | ML Forecast Methods | Drug Effects or Interventions |  |  |  | Continuous Intervention |  |  |  |
|  |  | 30-Min | 90-Min | 180-Min | 360-Min | 30-Min | 90-Min | 180-Min | 360-Min |
| Insulin Secretion ($a_1$) | LR | 0.90 ± 0.03 | 0.91 ± 0.03 | 0.93 ± 0.03 | 0.96 ± 0.03 | 0.84 ± 0.03 | 0.85 ± 0.03 | 0.85 ± 0.03 | 0.86 ± 0.03 |
| | SVR | 0.90 ± 0.08 | 0.91 ± 0.08 | 0.93 ± 0.08 | 0.95 ± 0.08 | 0.85 ± 0.08 | 0.85 ± 0.08 | 0.86 ± 0.08 | 0.86 ± 0.08 |
| | GPR | 0.92 ± 0.04 | 0.93 ± 0.04 | 0.95 ± 0.04 | 0.97 ± 0.03 | 0.87 ± 0.04 | 0.87 ± 0.04 | 0.88 ± 0.04 | 0.89 ± 0.03 |
| | LOESS | 2.59 ± 1.12 | 2.63 ± 1.17 | 2.70 ± 1.23 | 3.12 ± 1.64 | 2.60 ± 1.13 | 2.64 ± 1.17 | 2.68 ± 1.19 | 2.81 ± 1.29 |
| Insulin Clearance ($\tau_p$) (min) | LR | 0.77 ± 0.06 | 0.78 ± 0.05 | 0.81 ± 0.05 | 0.86 ± 0.05 | 0.76 ± 0.05 | 0.76 ± 0.05 | 0.77 ± 0.05 | 0.78 ± 0.05 |
| | SVR | 0.79 ± 0.15 | 0.80 ± 0.15 | 0.82 ± 0.15 | 0.86 ± 0.14 | 0.78 ± 0.14 | 0.78 ± 0.14 | 0.78 ± 0.14 | 0.79 ± 0.13 |
| | GPR | 0.86 ± 0.07 | 0.87 ± 0.06 | 0.89 ± 0.05 | 0.94 ± 0.04 | 0.85 ± 0.06 | 0.85 ± 0.06 | 0.85 ± 0.05 | 0.86 ± 0.03 |
| | LOESS | 3.14 ± 1.61 | 3.26 ± 1.71 | 3.58 ± 2.03 | 4.95 ± 3.40 | 2.93 ± 1.50 | 2.95 ± 1.53 | 3.18 ± 1.77 | 4.25 ± 2.88 |
| Insulin Resistance ($R_g$) (mg/min) | LR | 34.3 ± 4.19 | 34.6 ± 4.22 | 35.0 ± 4.20 | 35.9 ± 4.20 | 26.7 ± 3.83 | 26.7 ± 3.80 | 26.7 ± 3.81 | 26.7 ± 3.78 |
| | SVR | 37.1 ± 11.7 | 37.5 ± 11.7 | 38.0 ± 11.7 | 39.0 ± 11.8 | 29.6 ± 10.8 | 29.7 ± 10.9 | 29.7 ± 10.9 | 29.9 ± 11.0 |
| | GPR | 46.2 ± 3.82 | 46.6 ± 2.78 | 47.1 ± 2.11 | 48.1 ± 1.76 | 39.0 ± 3.40 | 39.1 ± 2.44 | 39.1 ± 1.92 | 39.4 ± 1.60 |
| | LOESS | 124 ± 76.9 | 129 ± 82.1 | 128 ± 81.7 | 141 ± 93.8 | 119 ± 75.8 | 120 ± 76.8 | 121 ± 78.2 | 123 ± 80.5 |

### 3.2 Sparse Real-World ICU Data Study

We demonstrate how our pipeline works for sparse clinical data by applying it to forecast the same underlying endocrine physiological parameters that were estimated in the simulation study.

This real-world evaluation roughly follows the simulation study, but in this case the ground truth of physiological changes does not exist. In contrast to the simulation study, we have no gold standard baseline with which to compare with because we are forecasting physiological mechanics, e.g., insulin secretion rate, that are typically unmeasured or unmeasureable in clinical settings. Meaning, there are no clinically available measurements that correspond to physiological parameters *a*_1_, *τ*_*p*_ and *R*_*g*_. While these physiological parameters are not directly observable, we performed external validations with HCP face validity analysis that confirmed the qualitative accuracy of these parameter estimates by means of physiological phenotypes in our prior work [12]. Additionally, because we are applying our previously validated DA machinery [12] to retrospective observational data, now we do have DA-based estimates for these mechanistic model parameters and treat them as ground truth. Furthermore, as our integrated pipeline forecasts these parameters estimated from retrospective observational data, we can calculate the MSE between the ML forecasts and the DA-based parameter estimates computed over the same time period as the forecasts. We will evaluate the ML forecast accuracy against these estimated parameters for all the ML methods.

#### 3.2.1 Clinical ICU Data

The physiological ODE model (cf. Sec. 2.1.1) requires inputs of glucose measurements and other observational data that correspond to modeled states in order to estimate model parameters (cf. Sec. 2.1.2). Clinical ICU data contain these data that include the glucose assays monitoring the general endocrine system state, the controlled nutrition and insulin therapies used as inputs, in addition to other EHR data external to the model such as demographics. The patient data set is comprised of ICU tube-fed patients from UCHealth Data Compass (Colorado Multiple IRB 18-2519). This data set includes intervals of 45 ICU stays associated with 45 patients who received short-acting insulin and had at least 20 glucose measurements within their last three days before extubation. These ICU stays are subintervals of encounters, where an ICU stay is a contiguous ICU period ([3,60] days) truncated by insertion and removal times of a feeding tube for regulated enteral nutrition.

Table 3 summarizes the inclusion criteria, nutrition and insulin therapy, and demographic information of the ICU stays in this tube-fed patient cohort restricted to the last three days before extubation. The longitudinal observational data from patient EHR records include: continuous enteral nutrition, non-bolus IV, and bolus IV deliveries; exogenous insulin therapies with continuous intravenous infusions, bolus intravenous injections, and subcutaneous injections; glucose measurements after the first non-zero nutrition in each ICU stay are used and these measurements are more frequent (every 1-2 hours) when patients receive continuous infusion insulin therapies rather than subcutaneous injections. The demographics of this patient cohort include: race, ethnicity, age, gender, length of stay, and number of insulin deliveries. Our previous work [12] includes information of another ICU cohort for mechanistic model parameters selection (cf. Sec. 2.1.3). This patient data set is comprised of nine ICU stays associated with nine ICU tube-fed patients from Columbia University IrvingMedical Center (Columbia University IRB AAAJ4503(M01Y06). The inclusion criteria, nutrition and insulin therapy, and demographic information of these ICU stays are similar to the details included in Table 3 . For other demographic information please refer to data set construction section in previous work [12].

**Table 3:**
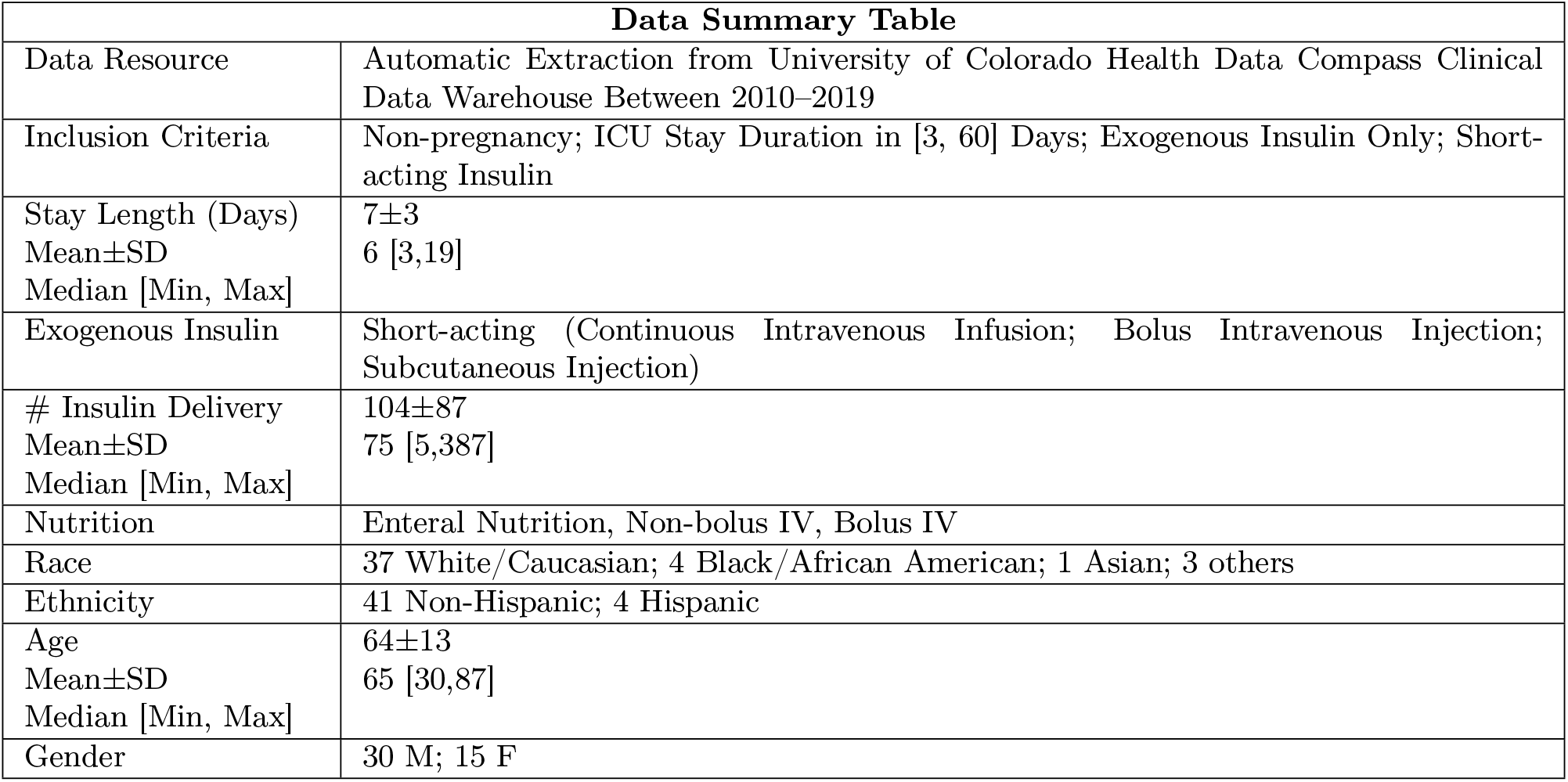
The nutrition and insulin therapy and demographic summary of 45 ICU stays, including data resources, data set inclusion criteria, ICU stay length in days, types of exogenous insulin therapy, insulin delivery counts, types of nutrition therapy, race, ethnicity, age, and gender. CGM data and insulin measurement are excluded, as is the general situation in an ICU setting. A large portion of patients is possibly on hemodialysis treatment.

| Data Summary Table |  |
| --- | --- |
| Data Resource | Automatic Extraction from University of Colorado Health Data Compass Clinical Data Warehouse Between 2010–2019 |
| Inclusion Criteria | Non-pregnancy; ICU Stay Duration in [3, 60] Days; Exogenous Insulin Only; Short-acting Insulin |
| Stay Length (Days)<br>Mean±SD<br>Median [Min, Max] | 7±3<br>6 [3,19] |
| Exogenous Insulin | Short-acting (Continuous Intravenous Infusion; Bolus Intravenous Injection; Subcutaneous Injection) |
| # Insulin Delivery<br>Mean±SD<br>Median [Min, Max] | 104±87<br>75 [5,387] |
| Nutrition | Enteral Nutrition, Non-bolus IV, Bolus IV |
| Race | 37 White/Caucasian; 4 Black/African American; 1 Asian; 3 others |
| Ethnicity | 41 Non-Hispanic; 4 Hispanic |
| Age<br>Mean±SD<br>Median [Min, Max] | 64±13<br>65 [30,87] |
| Gender | 30 M; 15 F |

#### 3.2.2 Generation of Longitudinal Data Sets

We use a time-ordered sequence of pre-specified data assimilation windows (DAW) to construct time trajectories of estimated physiological parameters for individual patients. Note that these parameter trajectories are estimated using observational data limited to each individual patient ICU stay (cf. Sec. 2.2). Parameter variability within each DAW is substantial because we do not borrow data across patients and clinical data are sparse, with only a few points per day. Therefore, to improve ML forecast robustness and reliability (cf. Sec. 2.3), we apply smoothing methods to overlapping DAWs to create smoothed analysis windows (SAWs) that are fed into ML methods as covariates. We estimate these SAWs by choosing the inference hyperparameter *a* while fixing explicit hyperparameters *b* = 1 and *δt* = 2 hours for an individual patient data. Figure 7 shows estimation of SAWs for an ICU stay of patient X results a range of *a* values (3, 18). The selected value of *a* = 6 balances trajectory temporal resolution and sufficient statistical representation (cf. Sec. 2.3.1), which defines the SAWs used for each of 45 ICU stays.

**Figure 7:**
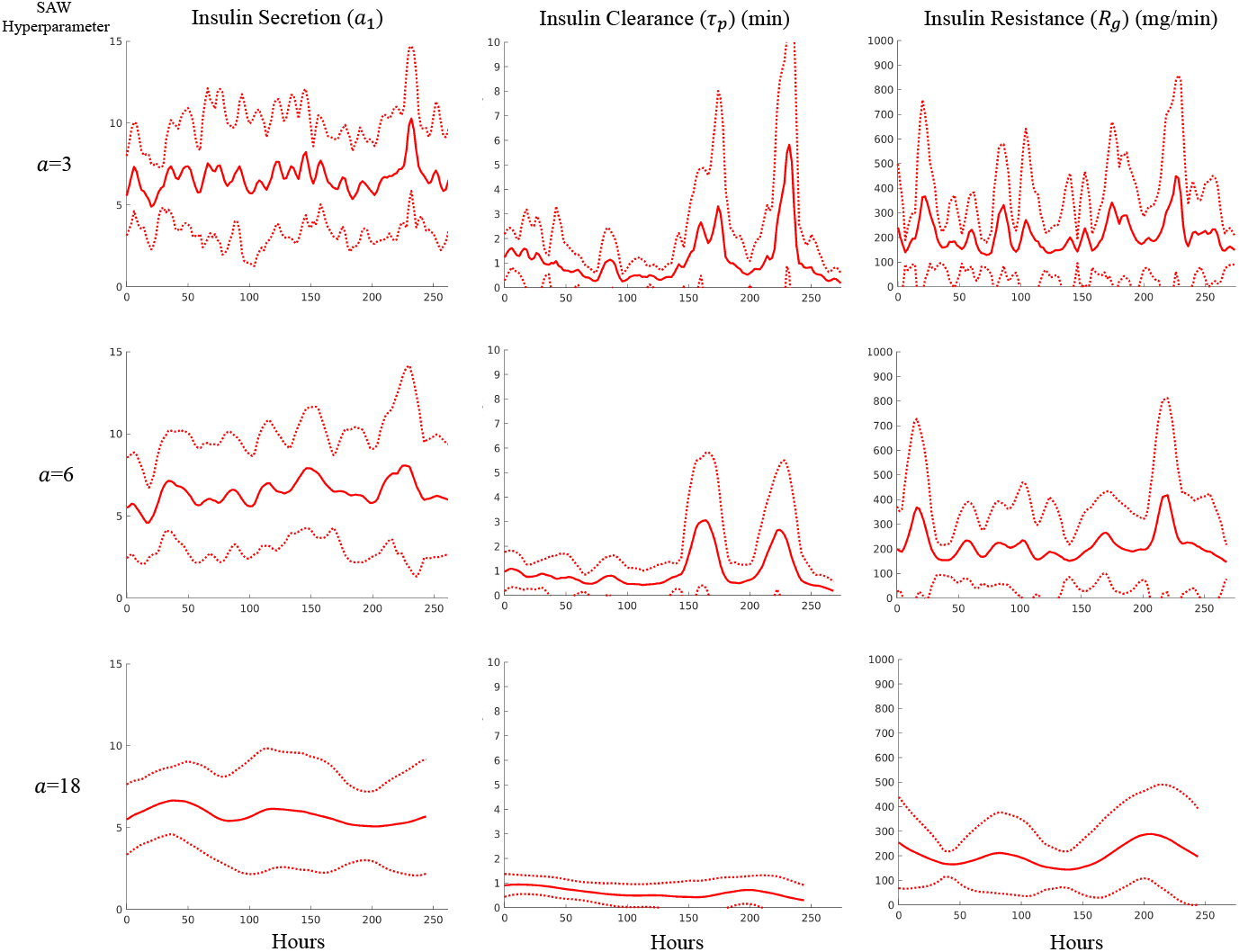
The impact of inference hyperparameter *a* in SAW estimation for constructing and smoothing parameterized physiological trajectory 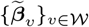 (mean *±* 2SD) for patient X. Hyperparameter *a* varies from 3, 6, to 18 across three physiological parameters (columns) to show the transition from high to low represented variability.

One possibility for boosting the data we have would be to include all patients together when computing physiological parameter forecasts. A potential problem with this approach is that real-world clinical data have inter-patient, intra-patient, and temporal heterogeneity. One source of temporal heterogeneity is the length of time patients are supported with tube feds, which is shown as the patient count curve in Figure 8 . This time length varies according to patient needs and leads to temporal heterogeneity across patients. For example, patients with longer tube feeding needs are generally more acutely ill. Moreover, there is substantial diversity in care delivery such as tube feed requirements. Figure 8 shows the population averaged smoothed parameter trajectory and individual patient smoothed parameter trajectories over the observed variable time patients were supported with enteral nutrition. It is worth noting that the patient count curve is not a survival curve of patients in ICU, but the number of patients with a given number of observations to estimate their physiological parameters at the time shown on the individual trajectories. For example, there are more patients with 50-hour than 45-hour long tube feeding encounters.

**Figure 8:**
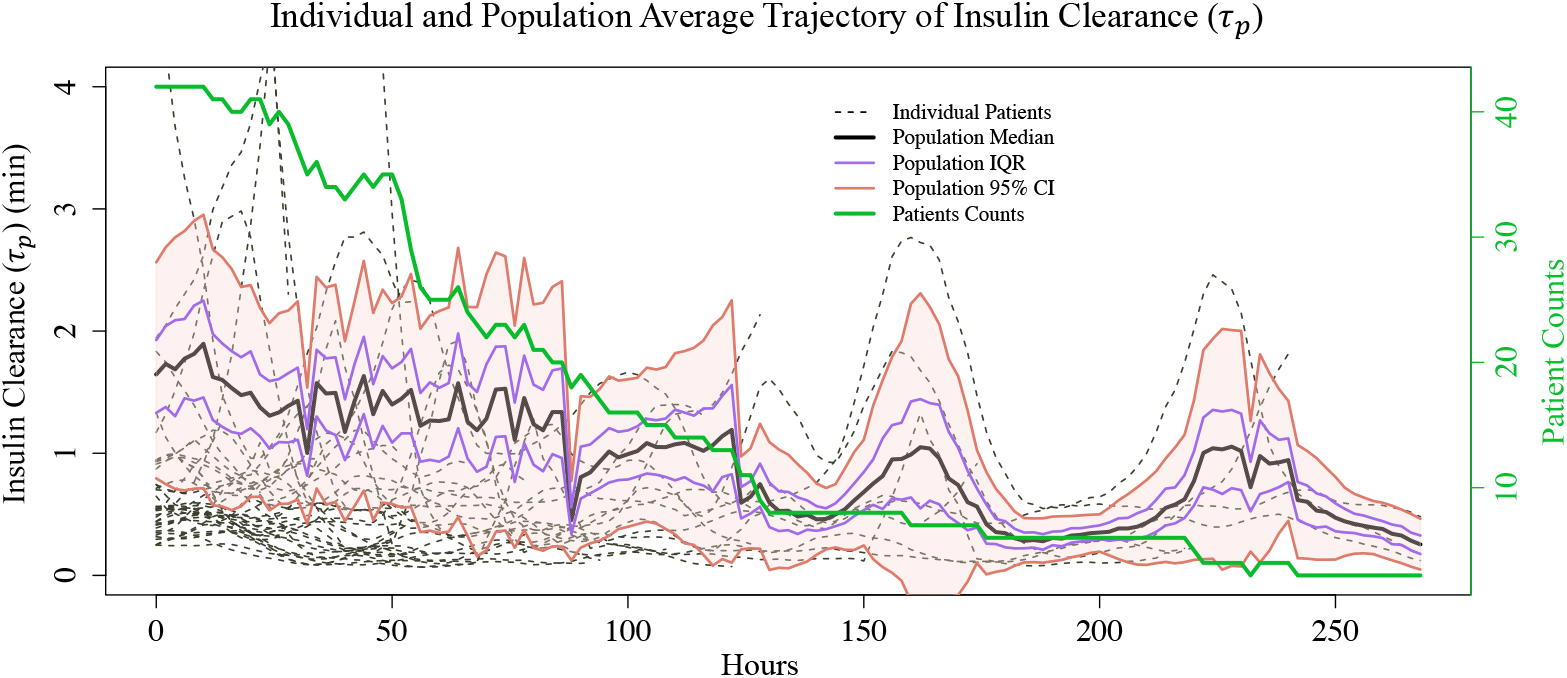
Smoothed parameter trajectory of insulin clearance (*τ*_*p*_) for 45 individual ICU stays (black dashed lines) and population average (solid lines) with median (black), IQR (purple), 95% CI of mean (orange) as patient counts (green) change over time. These patient counts represent the number of patients with a given number of observations to estimate their physiological parameters.

From Figure 8 we can observe inter-patient heterogeneity, variability in the length of time tube feeding is administered. We can also observe an important consequence of this temporal and inter-patient heterogeneity by observing how the variability and trend of the population averaged signal, here insulin clearance, changes over time as patients drop out of the population. Importantly, this signal generating process, or the characteristics of patient population, changes as the length of intubation increases, making the signal effectively non-stationary. Multiple peaks over the signal reveal the sensitivity to changes in fewer, longer-intubed patients, rather than changes in physiological mechanics. It is rather rare to have patients spend such long time in the ICU. In fact, more than 75% patients in the cohort are extubated before half of the length of the longest ICU stay, and (a) patients with short intubation times are rather different from patients with long intubation times, (b) there is no simple threshold or cutoff for intubation length that leaves two uniform sub-populations. These unique patients with longer stays differ from the general population clinically and likely in their parameter distributions of underlying physiological mechanics as well. Therefore, using population-averaged data can conceal pathophysiological processes unique to an individual ICU stay. This motivates constructing longitudinal ML data sets (cf. Sec. 2.4.1) based on individual-level physiological estimates for training and testing when possible. We will, however, compare the ML forecasts with individual patient data against a population-based method using functional data analysis.

#### 3.2.3 Qualitative Physiological Parameter Trajectory Evaluation

We use five ML forecasting methods to compute longitudinal regression forecasts of model parameters. These methods are trained and tested on the constructed longitudinal data sets comprised of the parameters estimated using DA. The ML methods hyperparameters are optimally tuned by hand. We pick these five methods because their different levels of flexibility to reconstruct and dynamically forecast the continuous parameter trajectory variations. Linear regression, support vector regression, Gaussian Process regression, and locally estimated scatterplot smoothing (LOESS) are estimated and evaluated using only an individual’s data. Functional concurrent regression, a filtered version of functional data analysis (FDA), is estimated on the population to account for intra-patient divergence. Notably, developing FDA methods that can be estimable using only an individual’s data is a topic of current research. Figure 9 shows forecasts of individual physiological progression at two forecast time points for three individual-based ML methods. Qualitatively, these methods all perform relatively well and support the necessary uncertainty quantification. Notably, LOESS and FDA are excluded due to their out-of-scale variability and inaccuracy.

**Figure 9:**
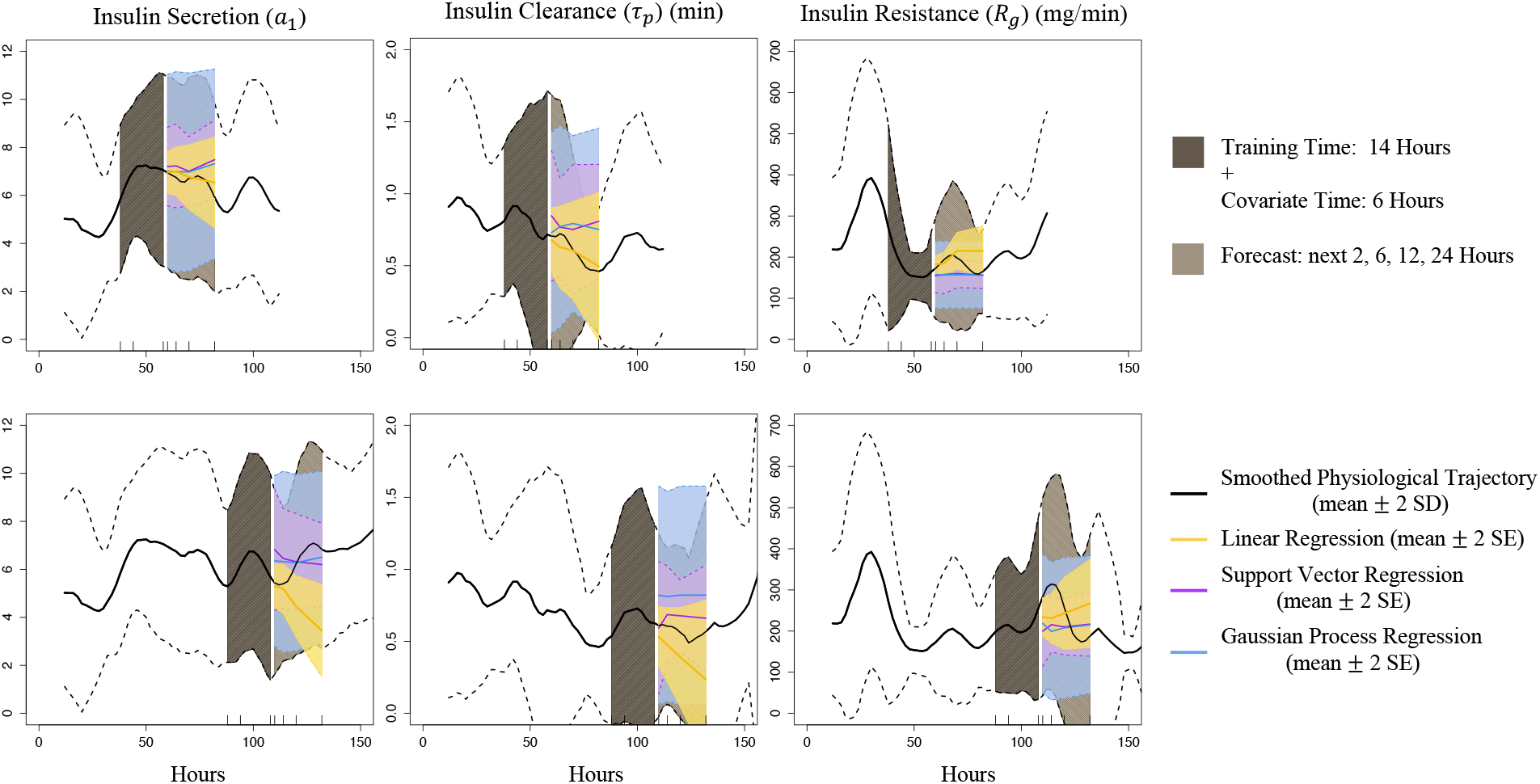
ML forecasting of individual physiological parameter at two time points (rows) in the next 2-24 hours using three ML methods, bounded by forecasts 95% CIs under normality assumption on top of smoothed physiological trajectory. These ML methods use the same historical physiological information (the past 14 hours for training with each data point containing covariates of its past 6 hours). Forecasts from LOESS are excluded due to relatively high variability.

#### 3.2.4 Quantitative Physiological Parameter Trajectory Evaluation

To quantitatively evaluate the pipeline we compare ML methods forecasts accuracy using the normalized root mean squared error (NRMSE) between the regression forecasts and data in the parameter space. We use the NRMSE, that is RMSE normalized over standard deviation, to eliminate the dependence of errors on parameter units. Table 4 shows the accuracy comparison in cohort-averaged NRMSE (*±* SE), partitioned by methods, covariate time, and training time. Linear regression has the overall lowest NRMSE for 2- and 6-hour forecasts. Support vector regression has the lowest NRMSE for longer forecasts over 12 and 24 hours. It is expected that linear methods minimize errors for short term forecasts, and interesting that as the time horizon increases, nonlinear methods are required to accurately forecast a given patient’s physiological progression. Population-trained functional data analysis has a higher NRMSE, likely due to inter-patient heterogeneity and the previously discussed impact of changes in the patient population for different time windows. Forecast accuracy consistently changes for all ML methods when the length of training time varies. However, increasing time windows over which the longitudinal regressions are trained does not always lead to a higher accuracy, likely due to inter- and intra-patient heterogeneity induced by the highly nonstationary nature of patient health trajectories in the ICU. Appendix Tables A.3 and A.4 show the bias and forecasts interval coverage probabilities using the estimated physiology as ground truth. Overall linear regression has the lowest bias, Gaussian process regression and functional data analysis have the highest coverage probability.

**Table 4:**
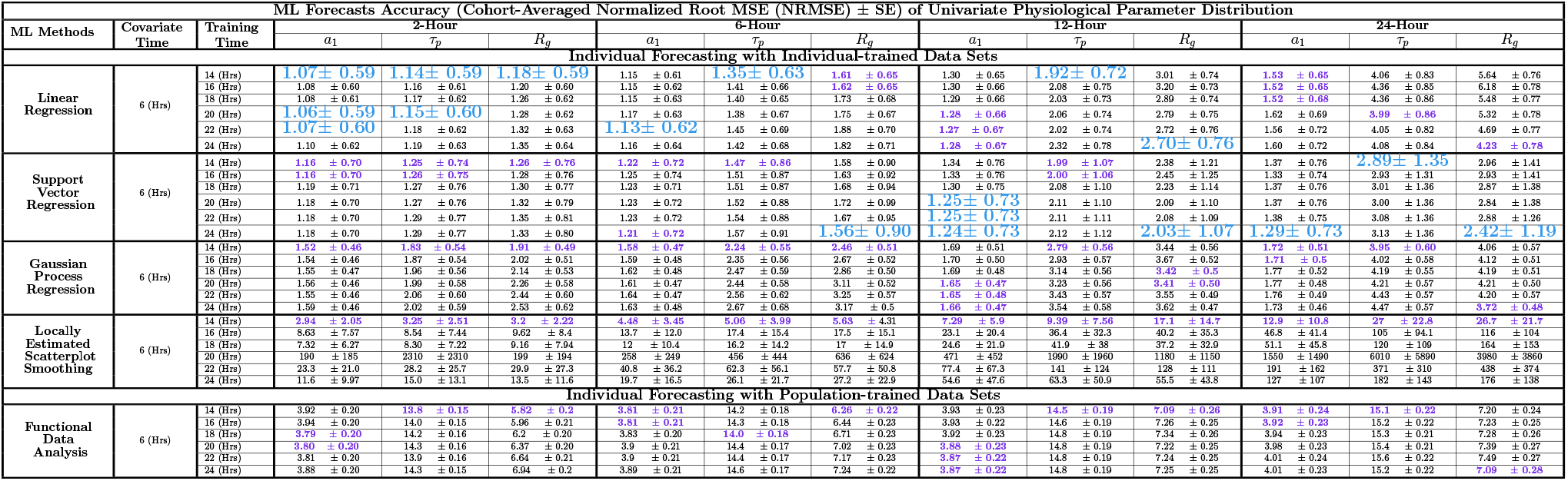
ML forecast accuracy of univariate physiological parameter distribution in the next 2, 6, 12, and 24 hours using normalized root MSE (NRMSE) averaged over cohort with uncertainty bounds (*±* SE). This table sectioned by five ML methods includes four individual-based methods and one population-based method. Each point in the training and testing data sets uses its previous 6-hour parameter information as covariates, where training time varies from 14 to 24 hours. This ML data set construction corresponds to using *P* = 3 and *k* − *j* ∈{7,, 12} (cf. Sec. 2.4.1). In each column within each ML method section, cells with the lowest average NRMSE are highlighted in color purple. Cells with the lowest values across all methods in the same column are colored blue. According to our UQ workflow (cf. Sec. 2.5), forecasting parameter distribution is based on 20 random samples (RS) when using linear regression, support vector regression, and locally estimated scatterplot smoothing methods. Forecasts are based on only 10 RS for faster computation when using Gaussian Process regression and functional data analysis.

## 4 Discussion

In this paper, we developed a new data assimilation (DA) and machine learning (ML) hybrid pipeline to estimate and forecast patient-specific trajectories of underlying physiological processes for individual patients using electronic health record (EHR) data. Specifically, this pipeline forecasts ordinary differential equation (ODE) models of physiological mechanics by forecasting model parameters of physiological processes. This integrated pipeline generates model-informed DA outputs that can be inserted into ML frameworks to address the clinical forecast problem and computational gaps. The computational pipeline development is the first step that sets the boundaries and limitations for future methods to work within, although the DA component is not the primary focus for making a contribution. This pipeline starts with DA model estimation that transforms sparse observational data into unobserved parameters of underlying endocrine processes in discrete data space. These parameters are then smoothed into continuous time series for ML training and forecasting. Some of the physiological processes we estimate, and forecast are not measured clinically. Because these true parameter values cannot be known, the parallel UQ workflow we developed is imperative for establishing reliability and clinical utility of parameter estimates and forecasts.

We validated and evaluated the hybrid forecast pipeline with simulation studies where we know the ground truth. This validation demonstrated that the pipeline accurately reconstructed and forecasted the pre-prescribed physiological parameter trajectories (cf. Sec. 3.1.2). We found that among four individual-trained ML forecasting methods, linear regression (LR) had the highest accuracy that minimizes root mean squared error (RMSE), while locally estimated scatterplot smoothing (LOESS) was the least accurate (cf. Sec. 3.1.3). We suspected the LR outperforms the nonlinear methods because these ML data are sparse, on the order of 10-20 data points per training and forecasting. Nevertheless, LR forecast changes in physiological mechanics when changes initially began to occur. Additionally, the kernel-based methods were not appreciably less accurate than linear regression. This interesting finding was consistent with standard approximation theory [46] that nonlinear models are more likely to outperform linear models when the system modeled are nonlinear and measured densely enough such that the nonlinearity can be resolved [47].

We also evaluated the pipeline with real-world clinical data. We applied the pipeline to individual tube-fed patients in ICU settings to forecast the same physiological parameters that were estimated in the simulation study. We used the pipeline to forecast univariate physiological parameters, e.g., insulin clearance, 2-24 hours into the future using an individual patient’s EHR data that are available operationally, usually between 12 and 15 data points. The performance of these ML methods was similar to the simulation studies.

Among the five ML forecasting methods, LR had the highest accuracy in normalized root mean squared error (NRMSE) for short term forecasting while support vector regression (SVR) had the highest accuracy over longer time scales (cf. Sec. 3.2.4). We suspect linear regression outperforms the nonlinear kernel methods because linear approximations are nearly always identical to or better than nonlinear forecasts [48, 49, 50, 46] for either short forecast horizons or when data are too sparse to contain much nonlinear information. In fact, short term changes are often effectively linear, while nonlinear changes tend to emerge over longer time scales. Our finding strikes a balance between nonlinear and linear modeling, because real-world clinical data sparsity renders many nonlinear regression methods less reliable for practical usage in processing nonlinear data. There are examples where nonlinear models can outperform linear models with similarly sparse data, but generally their nonlinear features are curtailed and primary gains come from UQ and robustness to outliers [51]. Population-trained functional data analysis (FDA) was less accurate than other individual-trained methods (except for LOESS). This lack of accuracy was likely due to a combination of intra-population diversity or heterogeneity and the relatively small ML data set. Population-estimated models such as FDA could be more reliable with more data and additionally provide useful insights into clinical diversity. Overall, these forecasts of physiological mechanics could substantially enrich available physiological information without requiring additional observational data by extracting and forecasting new computational biomarkers of unmeasurable endocrine mechanics. In a real-time prospective study, health care professionals (HCPs) could use these forecasts to, e.g., make decisions regarding insulin therapy.

### Simulation study is based on real-world data sparsity and patient non-stationarity

We designed our simulation study to parallel and mimic real-world data sparsity examples of non-stationary patients whose sampling processes and uniform time resolution are defined by clinical protocols beyond our control. While varying the sparsity in these observational data contributes to a methodologically meaningful evaluation in the simulation study, it is not directly translatable to a real data study with practical usage to demonstrate how the pipeline can support accurate forecasts. Nevertheless, to represent different responses to clinical interventions, we considered two curves as the ground truth of how the same underlying physiological process evolves differently using two different functions. The difference in generating model parameter time trajectories of these unobservable data reflects the heterogeneity of physiological non-stationarity that interacts with data sparsity simultaneously, both within and across patients. In the real data study, these processes have shared association with estimated model parameters whose changes are impacted by the same clinical decisions, leading to potentially similar but inherently different response curves.

### Uncertainty quantification (UQ) was embedded within workflow

To address the question of UQ when forecasting trajectory distributions, we introduced a UQ workflow with four stages (cf. Sec. 2.5). This workflow allows for both transparency and for UQ analysis of every step of our pipeline. The four stages of the UQ workflow include uncertainty identification and empirical approximation in UQ stage one, uncertainty management in UQ stage two and stage three, and uncertainty propagation with empirical approximation in UQ stage four.

### Extending UQ to both dependent and independent variables for reliable forecasting

Regardless of the ML method, an understanding of how empirical uncertainties in inputs determine resultant uncertainties in forecast estimates is critical for accurate and reliable forecasting. Generally speaking, only dependent variables are considered as source of uncertainties when they are modeled by methods with directional processes that entail an independent-variable to dependent-variable relationship [11]. Although this consideration is widely accepted, ignoring uncertainties of independent variables—here propagated uncertainties in UQ stage four—can lead to inconsistently biased coefficients or parameters inference [52], e.g., attenuation bias towards zero in linear regression. Here we demonstrated the necessity of generating multiple sets of random samples to construct ML data sets (cf. Sec. 2.5) to account for variability in the independent variables or measurements in the EHR. This construction involves random sampling historical estimates as training covariates in the design matrix (independent variables) and random sampling of current time estimates as training target in the response vector (dependent variables) (cf. Sec. 2.4).

### Alternative physiological interpretation of extended empirical UQ

We can interpret uncertainties of independent variables discussed above as physiologically present variability, as measurement error, or as a combination of both. Recall that each empirical random sample is generated from the parametric statistical distribution of smoothed physiological trajectories using Monte Carlo methods. This means that each random sample is an independent and idealized realization of the time evolution of these independent variables. This probabilistic setting allows for a continuum characterization of the independent variables, and we set the variability to reflect the randomness in observed physiological health changes over time [53] of actual patients. Therefore, we interpret uncertainties of independent variables predominantly as physiological variability to be propagated for clinical decision-making, instead of as measurements errors.

### 4.1 Limitations

#### Forecast performance could be affected by factors not accounted for in hybrid pipeline

According to Sec. 3.2.2, the intra-patient temporal variability implies sampling from individual patient rather than the population. In fact, heterogeneous physiological health [53] can be diversely impacted by interventions. However, without foreknowing insulin response to interventions, preceding information of estimated physiological processes could be overfit. While non-stationarity is managed by non-ML care-independent hyperparameters (cf. Sec. 3.2.2), this overfitting result could be causal to HCP role-dependent frequency factors, e.g., clinical shifts, that impact glycemic dynamics.

#### Idealized assumptions in current UQ workflow

Our Monte Carlo based UQ approach with the random sampling component was a solution chosen to ease pipeline implementation limited by computing resources (cf. Appendix A.4), while still enabling ML methods to provide forecast uncertainties with alternative biological interpretation justified above. We made three statistical assumptions in the current UQ workflow to improve efficiency and balance estimation quality with non-stationarity effects and data sparsity. However, these idealized assumptions complicate our UQ workflow between parametric and non-parametric forms. Different, less restrictive assumptions could provide similar and potentially less biased forecasts at higher computational costs. For example, we could skip the univariate normality assumption and instead estimate covariance that represents parameter synergy to improve forecast accuracy in a multivariate setting. Additionally, we assumed temporal independence between DAWs to generate ML covariates of individual processes. Even though we cannot eliminate the temporal dependence in the overlapping DAW-indexed estimates of continuous-time physiological processes, this assumption is unlikely to cause severe problems and practical for ML methods not targeting mixed effects or mean effect in the population. Furthermore, such data-and-temporal effects are localized over short timescales and their consequences could be reduced by alternate weighting schemes in future work.

### 4.2 Future work

#### Alternative modeling of extended empirical UQ

Theories have been developed by applied mathematicians and statisticians to treat empirical UQ, including symbolic data analysis that process ‘hypercube’ numerical data of histograms or intervals in the probabilistic metric space [54, 55, 56], and function-on-function methods in FDA that smooth quantile functions to forecast a distribution [57]. It is worth noting that forecasting distributions using complex nonlinear models, e.g., using SVR, solely based on numerical data still remains an active area of research.

#### Alternative UQ workflow

Many UQ options could otherwise render different results. *First*, using distribution divergence statistic, e.g., the Kullback-Leibler divergence, may improve optimization in estimation to facilitate subsequent smoothing processes. *Second*, we could generate random samples using stratified approaches [58, 59] that may require fewer statistical assumptions and cover the density space better than current methods depending on asymptotic distributions. One example is using distribution-independent approaches with parametric bootstrapping to propagate uncertainties, especially when care intervention decisions only rely on thresholds. *Third*, we currently used a fixed, manually selected stationarity hyperparameter *a* for all patients. The gain with a personalized or automatic optimization is minor given our methodology is already rather complex. Nevertheless, an optimized criteria for each individual patient is a potential future direction of research. *Fourth*, parameter smoothing was based on the temporal intersection of overlapping windows, a methodological choice that reduced temporal dependence of estimates while implicitly incorporating the influence of observation density, e.g, number of data points, on these estimates into downstream uncertainty. Further dependency reduction and explicit consideration of observation density in the UQ pipeline are important future developments.

#### Methodological extensions

In the current work, ML forecasts only included DA outputs. One potential next step would be to include external variables, e.g., line of therapy or real-valued drug dosages, in the longitudinal forecasting pipeline. These extensions may lead to the development and use of deep learning methods, e.g., neural ODE, and of nonparametric models to forecast the same physiological processes.

#### Generalizability

Developed for tube-fed ICU patients potentially on IV insulin, this pipeline could be generalized to non-ICU or non-tube-fed patients in the current modeling context [60] or to other physiological contexts [61, 62].

## Supporting information

Supplementary animated videos for section 3.1.2

## 5 Data Availability

Due to HIPAA compliance, the ICU patient data that have been used in this work would remain confidential and would not be shared. This pipeline is developed in the Matlab and R environment, and the source code can be shared upon request.

## 6 Funding Acknowledgments

The work was supported by grants from the National Institutes of Health R01 LM006910 “Discovering and applying knowledge in clinical databases,” and LM012734 “Mechanistic machine learning,”.

### A

#### A.1 The Ultradian Glucose-Insulin ODE Model

The mechanistic ultradian model [20] in the form of mathematical formulas has six ordinary differential equations with three physiologic states (*I*_*p*_ is plasma insulin, *I*_*i*_ is remote insulin, *G* is plasma glucose) and a three stage filter (*h*_1_, *h*_2_, *h*_3_) that represents how plasma insulin affects glucose. Below shows model equations and its full list of 21 parameters.

| Full List of Model Parameters |  |
| --- | --- |
| Parameter | Nominal Value (Unit) |
| $V_p$ | 3 (l) |
| $V_i$ | 11 (l) |
| $V_g$ | 10 (l) |
| $E$ | 0.2 (l min<sup>-1</sup>) |
| $\tau_p$ | 6 (min) |
| $t_i$ | 100 (min) |
| $t_d$ | 12 (min) |
| $k$ | 0.5 (min<sup>-1</sup>) |
| $R_m$ | 209 (mU min<sup>-1</sup>) |
| $a_1$ | 6.6 |
| $C_1$ | 300 (mg l<sup>-1</sup>) |
| $C_2$ | 144 (mg l<sup>-1</sup>) |
| $C_3$ | 100 (mg l<sup>-1</sup>) |
| $C_4$ | 80 (mU l<sup>-1</sup>) |
| $C_5$ | 26 (mU l<sup>-1</sup>) |
| $U_b$ | 72 (mg min<sup>-1</sup>) |
| $U_0$ | 4 (mg min<sup>-1</sup>) |
| $U_m$ | 94 (mg min<sup>-1</sup>) |
| $R_g$ | 180 (mg min<sup>-1</sup>) |
| $\alpha$ | 7.5 |
| $\beta$ | 1.772 |

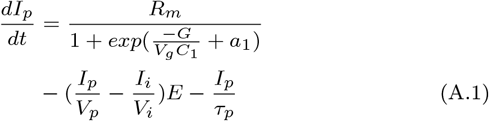

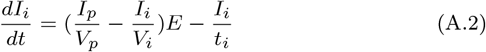

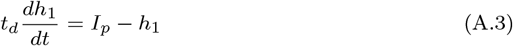

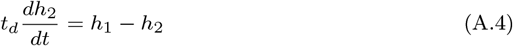

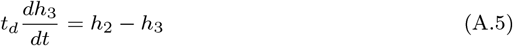

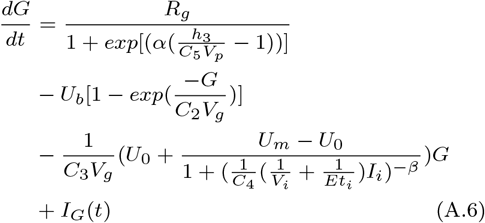

#### A.2 Mathematical Notations for Constructing ML Data Sets

To better mathematically present the structure of design matrix 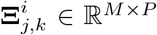, we make the following simplification notes by defining three operators on a vector *x* = (*x*_1_, *x*_2_, · · ·, *x*_*W*_ ): (i) a left-shift operator ℒ with ℒ_1_(*x*) = (0, *x*_1_, *x*_2_, · · ·, *x*_*W* −1_) that adds zero to the empty space on the left such that ℒ_*m*_(*x*) = (0, …, 0, *x*_1_, *x*_2_, …, *x*_*W* −*m*_) is the transformed vector by applying the left shift operator *m* times. This operator is analogous to the Box-Jenkins backshift operator [63, 64] that simplifies a autoregressive moving averages model, (ii) a similar right shift operator ℛ with ℛ_1_(*x*) = (*x*_2_, · · ·, *x*_*W*_, 0), (iii) a clip operator *C*_[*i,j*]_ that keeps a few elements between indices *i* and *j* with 1 ≤ *i < j* ≤ *W*, where *C*_[*i,i*]_(*x*) = (*x*_*i*_) and *C*_[*i,j*]_(*x*) = (*x*_*i*_, *x*_*i*+1_, · · ·, *x*_*j*_). Using operators ℒ, ℛ, *C* defined above, the mathematical operation ℝ^*W*^ ↦ ℝ^*M ×P*^ to construct 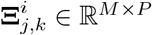 with any *i, j* is written as

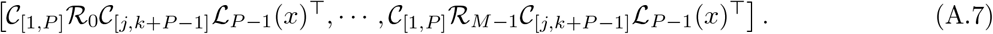

Depending on ML method to handle potential multicollinearity, the exact *x* used in practical implementation to generate 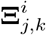 is one of the following vectors: (i) SAW indices (1, 2, · · ·, *W* ), (ii) one-dimensional physiological parameter component 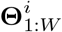, and (iii) a concatenation of (i) and (ii) by data columns.

#### A.3 Specific Forms of Forecasting Machineries

##### A.3.1 Support Vector Regression (SVR)

When we use SVR [65, 66] to forecast univariate physiological parameter of an individual patient *i*, we have *h* in the specific form with *< *, * >* being the dot product

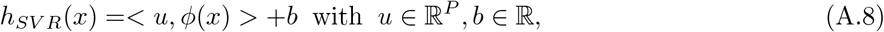

which also specifies SVR hyperparameters *λ*_*ML*_ = {*u, b*}. Vector *x* represents a feature row and ϕ(*x*) is its SVR projection into high dimensional space ϕ : ℝ^*P*^ ↦ F. The optimal *u*_*opt*_ is found via either minimization of a primal problem or a maximization of a dual problem given training set 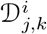 with design matrix 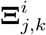 and response target vector 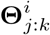, such that 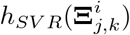 is within a short interval near 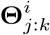 . The optimal *b*_*opt*_ is calculated under the Karush-Kuhn-Tucker conditions. To forecast parameter in future time interval *S*_*v ′*_ using the feature vector 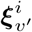, we then have

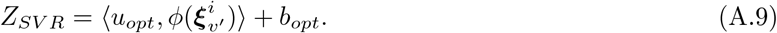

##### A.3.2 Gaussian Process Regression (GPR)

Gaussian Process Regression (GPR) [67] is an application of Gaussian Process (GP) that projects input vector *x* into a set of basis functions in high dimensional space. In our physiological trajectory forecasting context, we describe a GP as a collection of random variables *g*(*x*) that are indexed by discrete SAWs and satisfy the Gaussian property, finite variance, and stationarity. We take additive Gaussian errors into consideration for training output function *h*_*GP R*_(*x*)

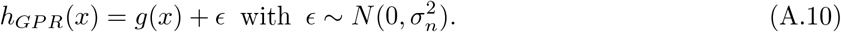

The fundamental idea behind GPR, as described by Rasmussen [67], is the representation theorem, or the kernel trick. This theorem from a Bayesian function-space view represents the covariance function *K* of *g*(*x*) as an inner product of *x* in lower dimensional input space. Under the representation theorem, our decision procedure is to find the optimal action of GPR hyperparameters *λ*_*ML*_ = {*l, σ*_*h*_, *σ*_*n*_} that define the penalty for making an inaccurate forecast. Here *l* is the process length-scale, 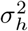 the signal variance, and 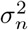 the noise variance of the prior covariance function *K*. For each individual patient *i*, we set our optimization decision as a Bayes risk function that uses a squared loss function, where the optimal action is the minimum contrast estimate of the Bayes risk conditional on observation function 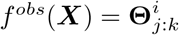 with ***X*** coming from rows of design matrix 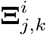. For a future time interval *S*_*v*_′, forecasts using the optimal *λ*_*ML*_ are sampled from the predictive posterior distribution

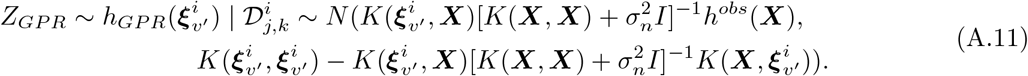

##### A.3.3 Locally Estimated Scatterplot Smoothing (LOESS)

LOESS [68] aims to find the best locally fitted smooth curve using lines and parabolas that obtain the lowest weighted residual sum of squares (RSS) in the training set 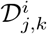 . In a local linear fitting case using matrix *λ*_*ML*_ = *A*, for each individual patient *i*, we denote the RSS

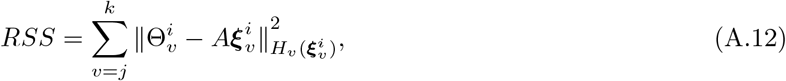

where each 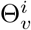 is a scalar target response, 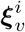 is a training feature vector, and local weight function 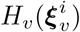 is a symmetric, positive-definite matrix of metric on the target space. For forecasting at the future test SAW *v′* using optimal *A*_*opt*_, we then have

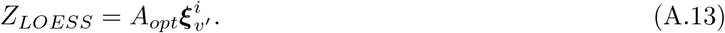

##### A.3.4 Functional Data Analysis (FDA)

Compared to above three ML algorithms that are trained on individual patients, we use FDA that is trained on the cohort to forecast individual trajectory. Specifically, we use the functional concurrent regression (FCR) [57] that in a continuous functional view trains on a cohort of physiological trajectories to forecast progression in individual patient. The FCR incorporates patient-specific random effects into the modeling process to set up spline basis. Using cohort training data set that is the row concatenation of 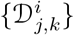 by patients, the general form of this machinery for patient *i* is

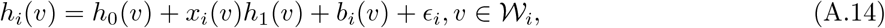

where *v* is the SAW index representing some aspects of time. Here *h*_0_(*v*) is the population intercept function, *x*_*i*_(*v*) represents patient-specific design matrix 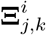 in the training set 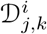, *h*_1_(*v*) is the population-fitted concurrent association of *x*_*i*_(*v*) with a population of patient-specific response functions of training values 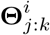, the Gaussian Process *b*_*i*_(*v*) ∼ *GP* (0, *C*(*v, v′*)) is patient-specific random functional deviation from the mean with *C*(*v, v′*) being approximated by basis functions {*B*(*v*)} through decomposition, and error term 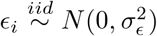. We have 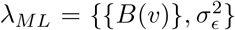 optimized to give the best linear unbiased predictor (BLUP) estimates. For individual patient *i*, we then denote forecast for the future in time interval *S*_*v′*_ as

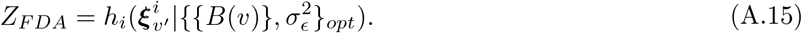

This cohort-wide training approach can forecast individual trajectory at unobserved time points that are still in-sample relative to the cohort. A reader interested in its specific form is referred to [69].

#### A.4 Resolving Pipeline Implementation Complexity at Cohort Level

To deal with information redundancy in high dimensions from MCMC sampling, we scale down a cohort of MCMC-inferred personalized estimates of three physiological parameters before storing them on disk for faster cohort-level implementation. We compress sample data of each parameter to its empirical mean, variance, and IQR, assuming no parameter synergy, univariate normality, and independence between estimation time before running the pipeline. This empirical density compression massively lowers data storage requirement from 45G to only 10KB for estimating all 45 ICU stays in the cohort. Resembling pipeline step four, we then randomly sample data from a univariate normal distribution parameterized by these compressed empirical information to represent the original uncertainties in estimates. Assuming across-patient independence, we use the cohort-averaged empirical mean and cohort-averaged total empirical variance summed over all patients to represent cohort-level uncertainties based on characteristic functions in probability theory.

### A.5 Simulation Study Additional Tables

**Table A.1:**
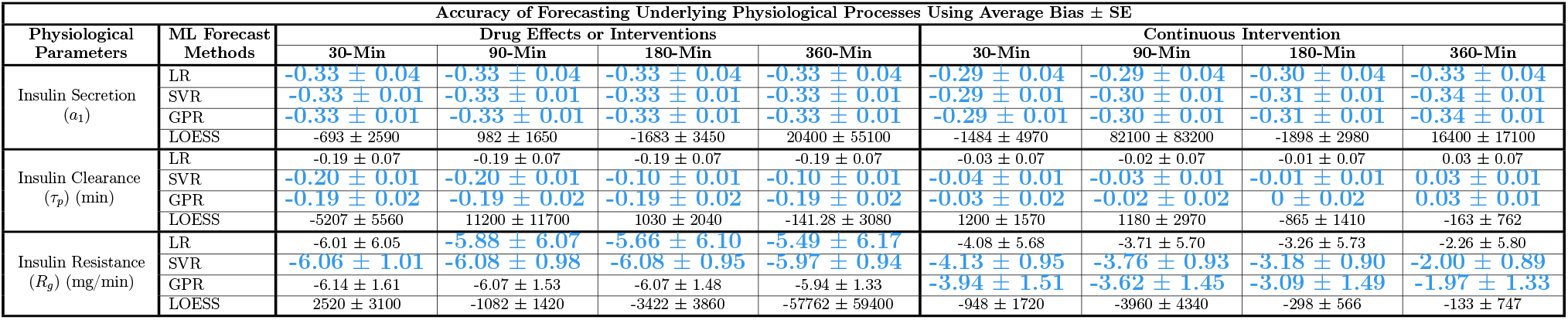
Evaluation of ML forecasting univariate physiological parameter distributions using bias averaged over 10 simulations with uncertainty bounds (*±* SE). This evaluation process is separated by experiments assuming insulin response that are (A) rapid and immediate, or (B) slower with onset and decay continuously, with forecast accuracy of parameters (row sections) at four future time points (columns). For each parameter, cells highlighted in color blue have the lowest average bias among four ML methods. According to our UQ approach (cf. Sec. 2.5), forecasting parameter distribution is based on 20 random samples (RS) when using linear regression (LR), support vector regression (SVR), and locally estimated scatterplot smoothing (LOESS). Forecasts are based on 10 RS for faster computation when using Gaussian Process regression (GPR).

**Table A.2:**
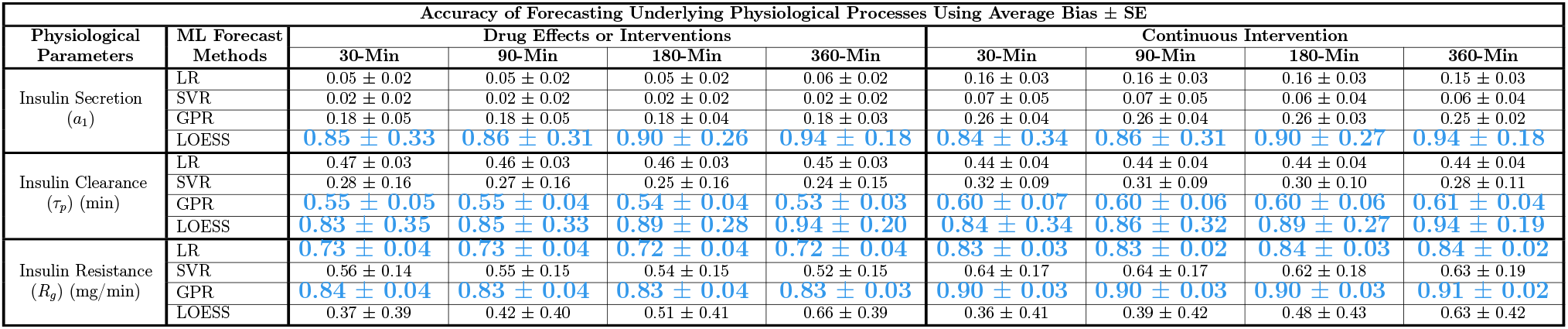
Evaluation of ML forecasting univariate physiological parameter distributions using coverage probability averaged over 10 simulations with uncertainty bounds (*±* SE). This evaluation process is separated by experiments assuming insulin response that are (A) rapid and immediate, or (B) slower with onset and decay continuously, with forecast accuracy of parameters (row sections) at four future time points (columns). For each parameter, cells highlighted in color blue have the highest average coverage probability among four ML methods. According to our UQ approach (cf. Sec. 2.5), forecasting parameter distribution is based on 20 random samples (RS) when using linear regression (LR), support vector regression (SVR), and locally estimated scatterplot smoothing (LOESS). Forecasts are based on 10 RS for faster computation when using Gaussian Process regression (GPR).

#### A.6 Sparse Real-World ICU Data Study Additional Tables

**Table A.3:**
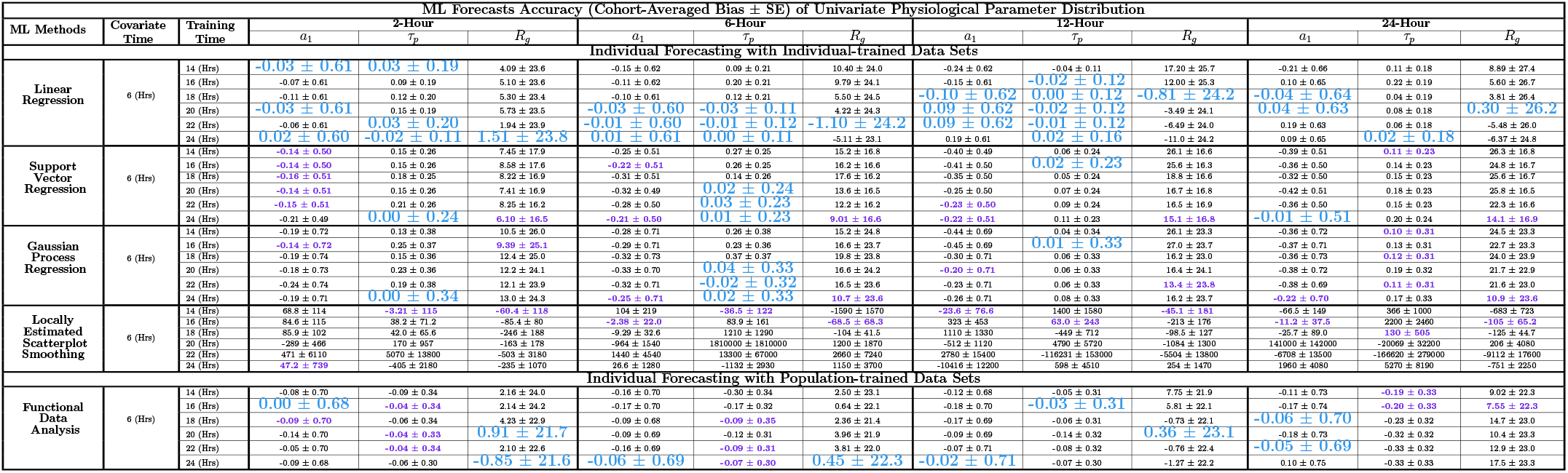
ML forecast accuracy of univariate physiological parameter distribution in the next 2, 6, 12, and 24 hours using bias averaged over cohort with uncertainty bounds (*±* SE). This table sectioned by five ML methods includes four individual-based methods and one population-based method. Each point in the training and testing data sets uses its previous 6-hour parameter information as covariates, where training time varies from 14 to 24 hours. This ML data set construction corresponds to using *P* = 3 and *k* − *j* ∈ {7, *…*, 12} (cf. Sec. 2.4.1). In each column within each ML method section, cells with the lowest average bias are highlighted in color purple. Cells with the lowest values across all methods in the same column are colored blue. According to our UQ workflow (cf. Sec. 2.5), forecasting parameter distribution is based on 20 random samples (RS) when using linear regression, support vector regression, and locally estimated scatterplot smoothing methods. Forecasts are based on only 10 RS for faster computation when using Gaussian Process regression and functional data analysis.

**Table A.4:**
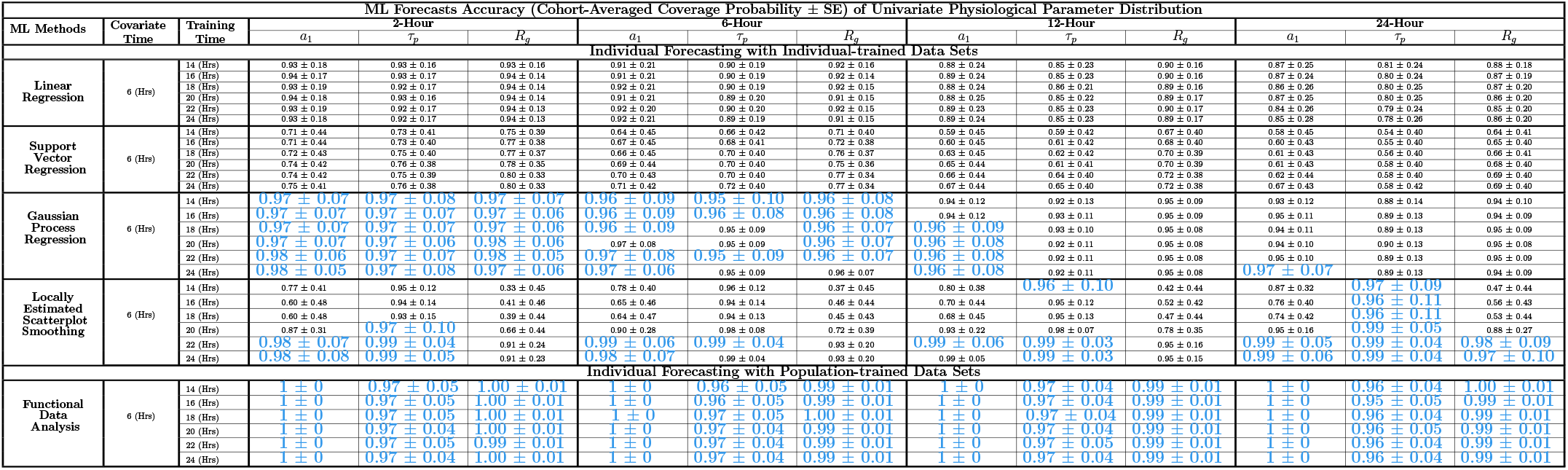
ML forecast precision of univariate physiological parameter distribution in the next 2, 6, 12, and 24 hours using coverage probability averaged over cohort with uncertainty bounds (*±* SE). This table sectioned by five ML methods includes four individual-based methods and one population-based method. Each point in the training and testing data sets uses its previous 6-hour parameter information as covariates, where training time varies from 14 to 24 hours. This ML data set construction corresponds to using *P* = 3 and *k* − *j* ∈ {7, · · ·, 12} (cf. Sec. 2.4.1). Cells with the highest values across all methods in the same column are colored blue. According to our UQ workflow (cf. Sec. 2.5), forecasting parameter distribution is based on 20 random samples (RS) when using linear regression, support vector regression, and locally estimated scatterplot smoothing methods. Forecasts are based on only 10 RS for faster computation when using Gaussian Process regression and functional data analysis.

