## Supplementary figures and images for "Forecasting Trajectories of Physiological Mechanics with Sparse Clinical Data Using a Data Assimilation and Machine Learning Hybrid"

### Figure6_Experiment(A)_a1.gif

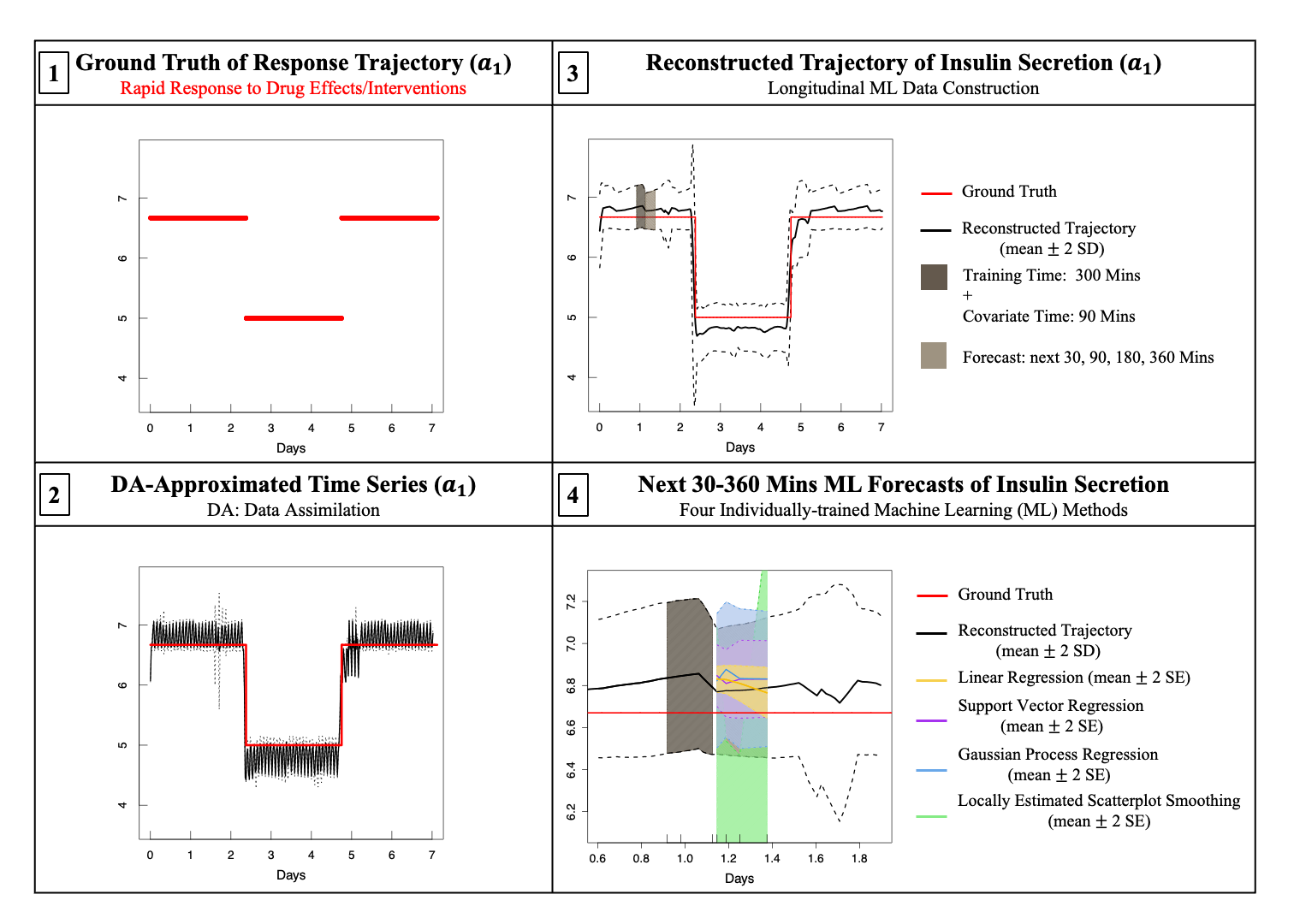

### Figure6_Experiment(A)_Rg.gif

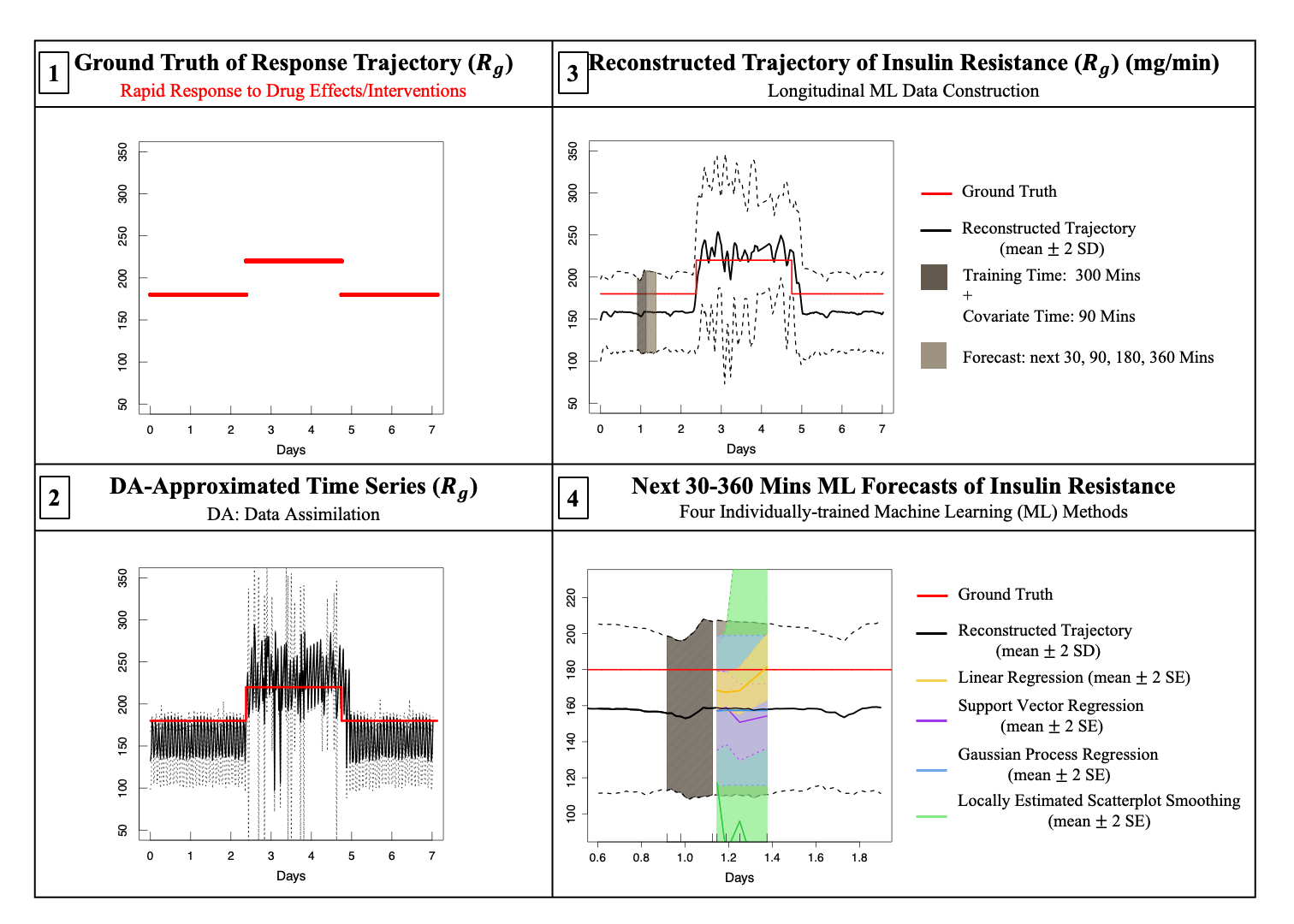

### Figure6_Experiment(A)_tp.gif

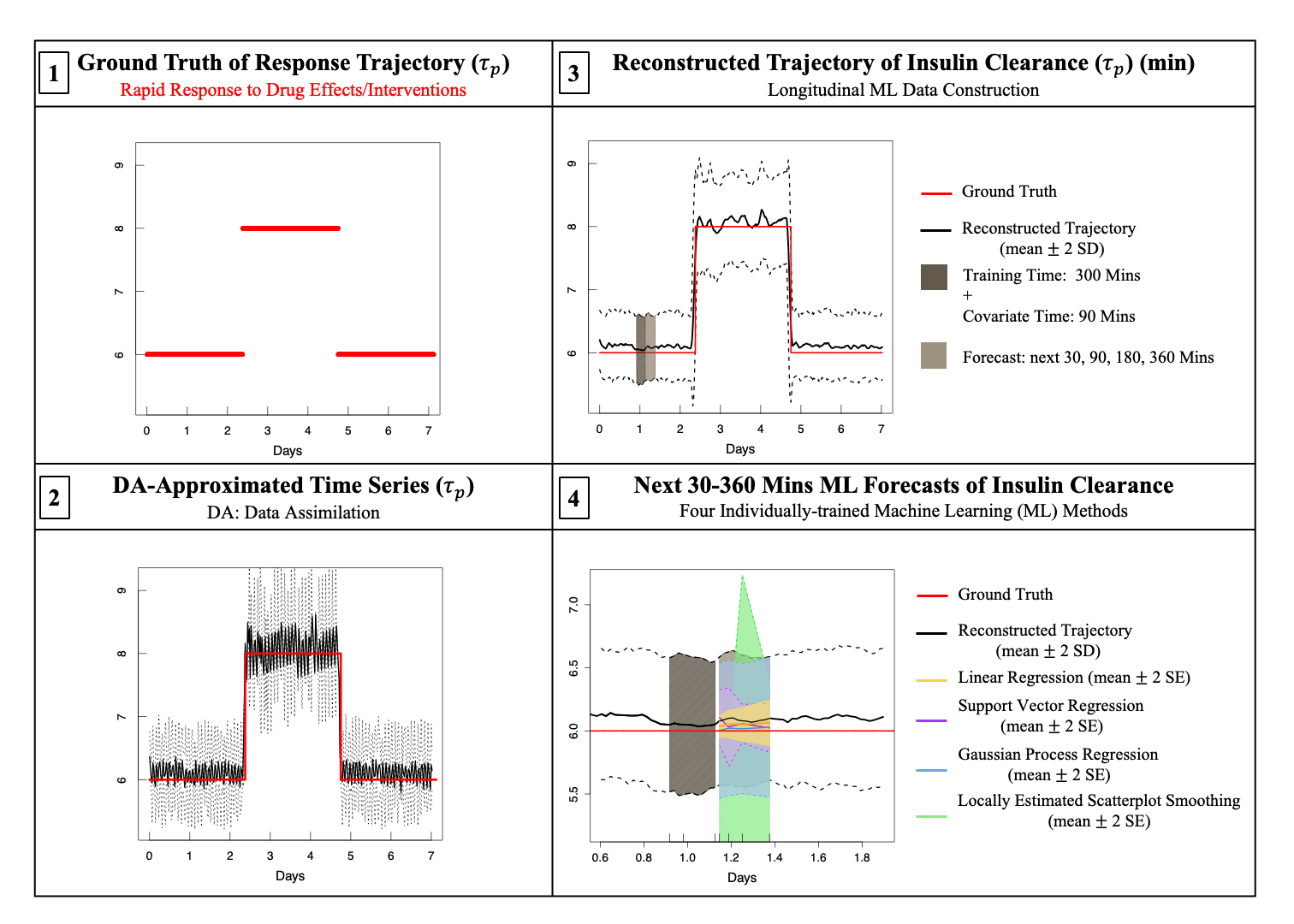

### Figure6_Experiment(B)_a1.gif

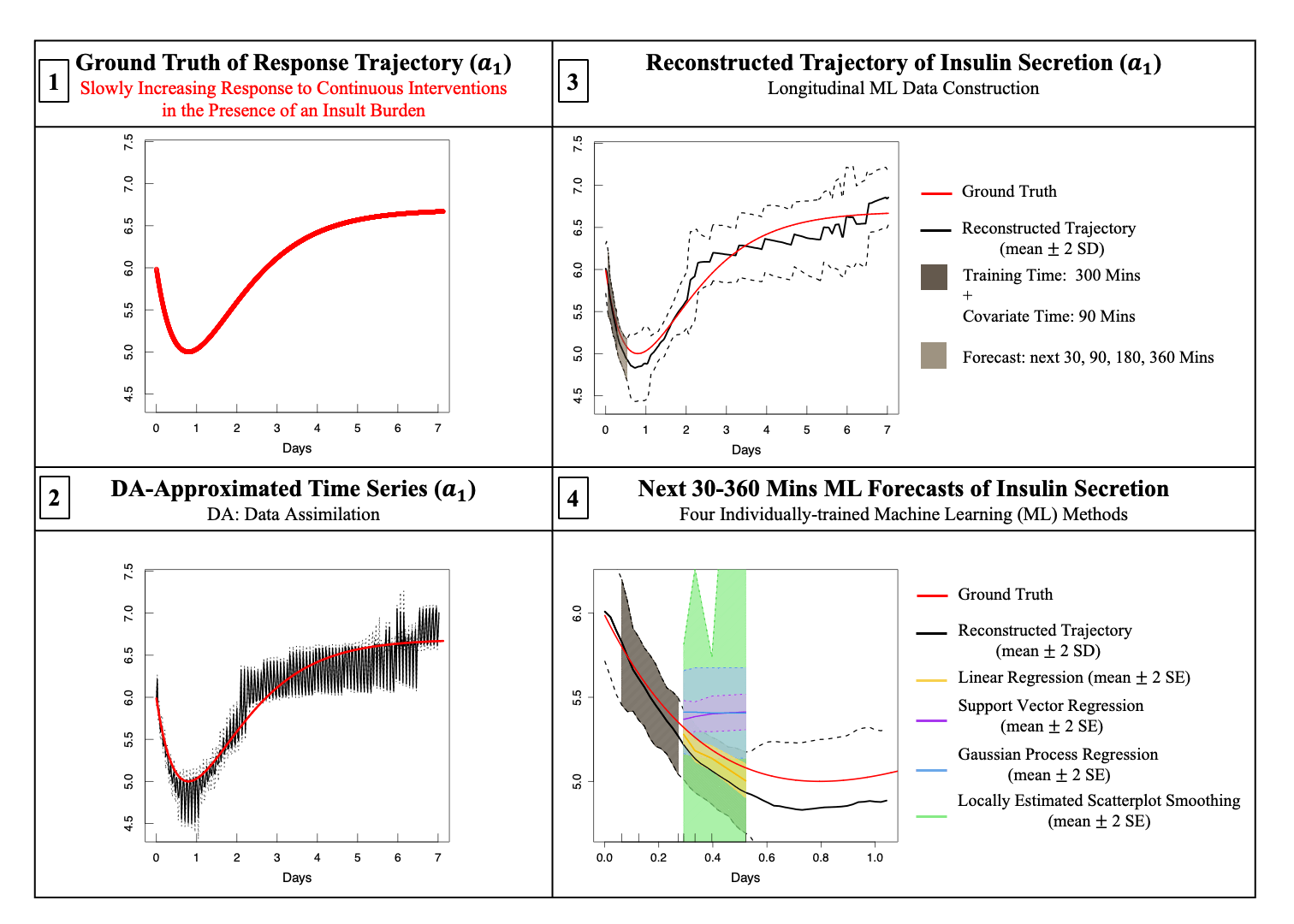

### Figure6_Experiment(B)_Rg.gif

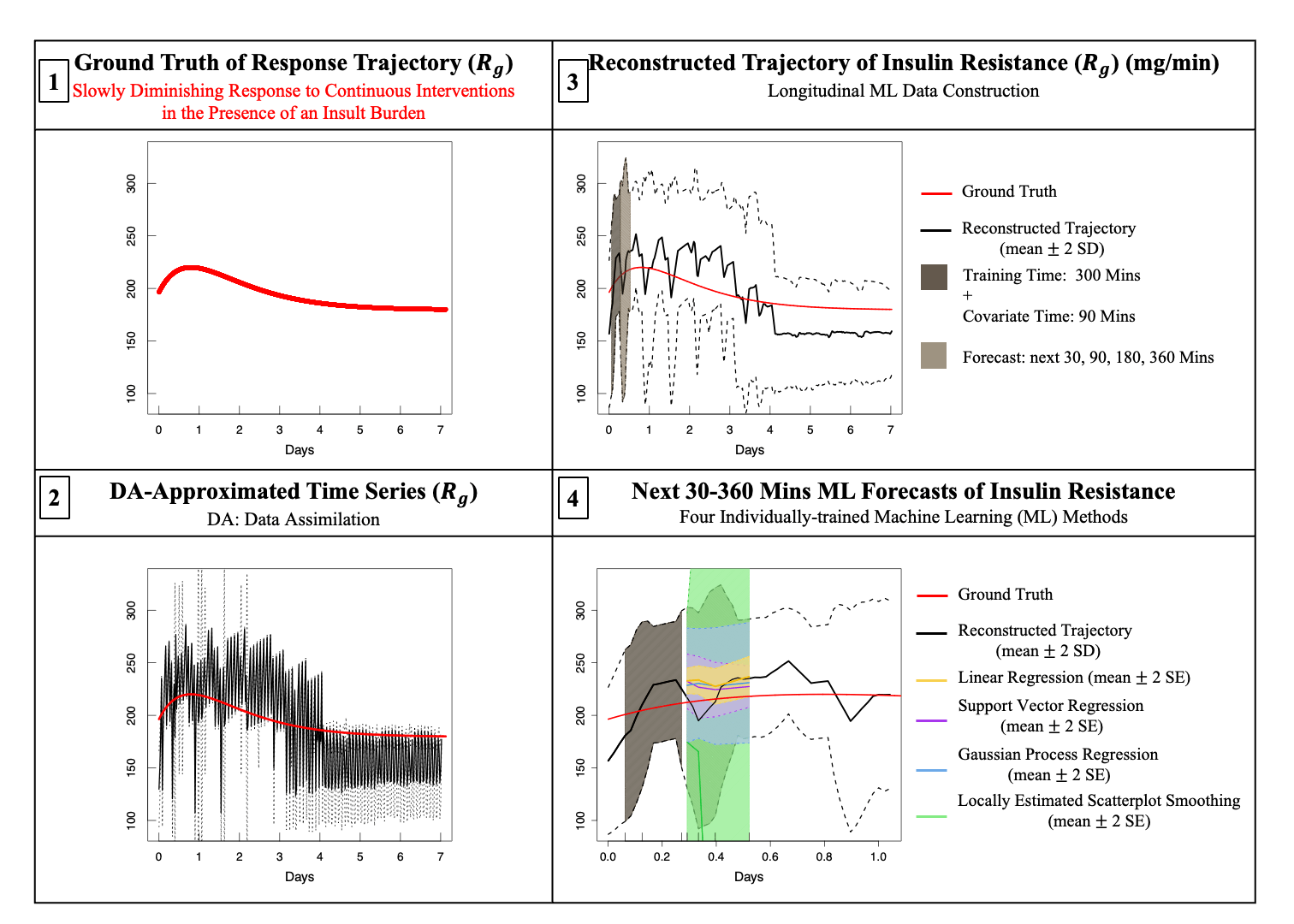

### Figure6_Experiment(B)_tp.gif

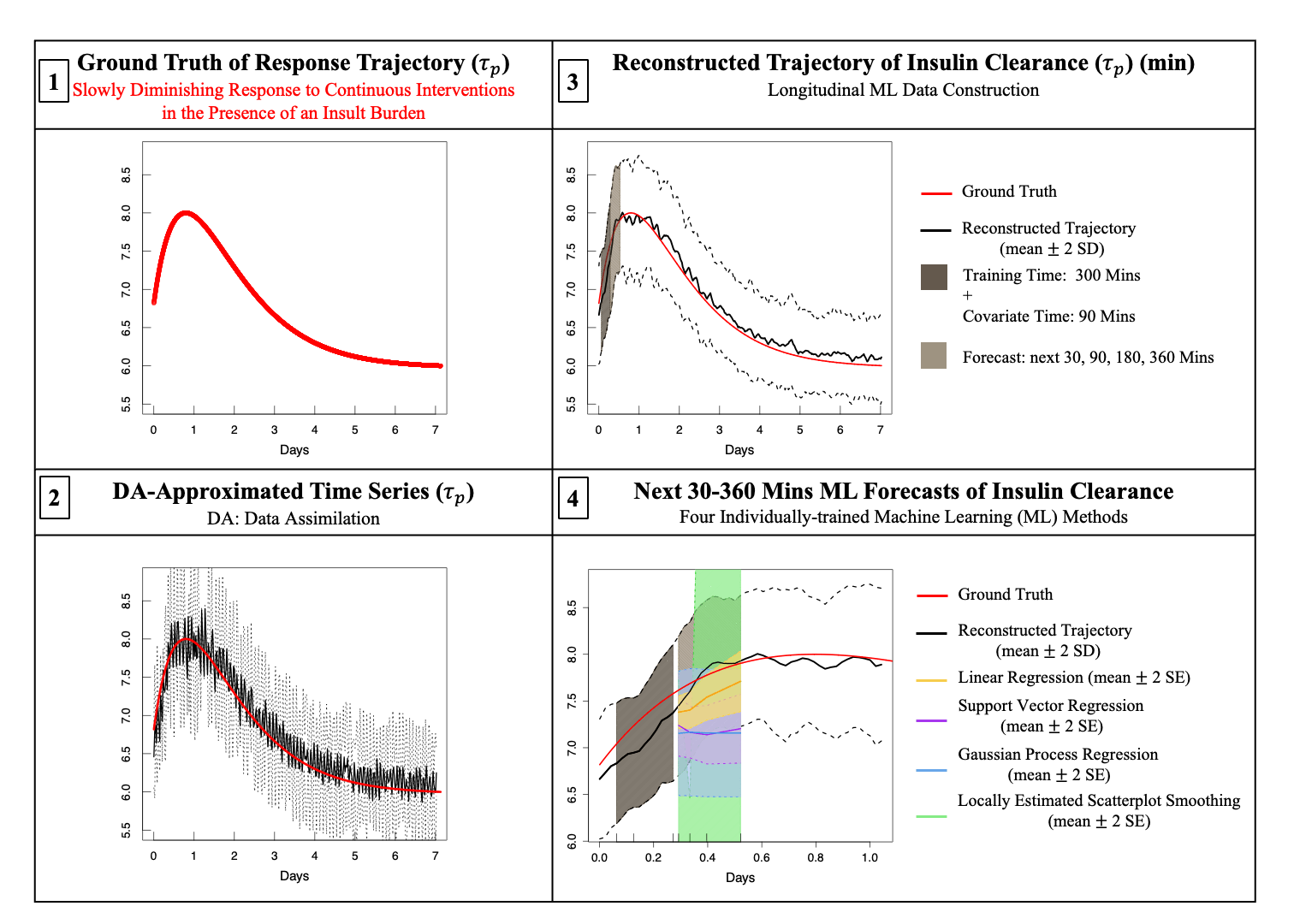
